# LDSC++: Improving linkage disequilibrium score regression estimation of heritability and genetic correlation for multivariate GWAS analysis

**DOI:** 10.1101/2025.04.07.25324946

**Authors:** Johan Zvrskovec, Alexandra Gillett, Michel Nivard, Jonathan R I Coleman, Christopher Hübel, Raquel Iniesta, Gerome Breen

## Abstract

**Introduction:** Linkage disequilibrium (LD) score regression is widely used for estimating common variant heritability and genetic correlations from genome-wide association study (GWAS) summary statistics. We hypothesise that segmented regression (also known as piecewise regression) improves on previous LD score regression implementations, when estimating both genetic covariance and its standard error.

**Methods:** We present novel extensions to LD score regression (LDSC++) improving I.) handling of varying numbers of shared genetic variants across trait pairs and reference panels, II.) estimation of genetic covariance and its variance, and III.) handling of imputation quality. We propose supporting statistical tests that use our novel extensions to improve sensitivity, and are further aimed at comparing parameter estimates that are highly correlated, such as those obtained from the same trait but from different methods. We validate LDSC++ first on real-world individual level data from the Genetic Links to Anxiety and Depression study and the United Kingdom National Institute of Health and Social Care Research BioResource (N: 14,190 - 20,144), second on simulated data with different degrees of shared QTL, and third on a battery of publicly available GWASs of ten diverse traits of varying statistical power and heritability.

**Results:** Using variance-component method (GCTA-GREML) estimates for reference, LDSC++ extensions were found to yield heritability estimates with a bias of about -10% to -20% while standard LD score regression yielded a bias of -30%, and heritability variability estimates with a bias of -1% to -7% while standard LD score regression yielded a bias of 8%. For ten external trait GWASs, LDSC++ was shown to recover 5% to 8% larger heritabilities with 4% smaller variability on average compared to standard LD score regression. Weighting by imputation quality in the model, rather than excluding genetic variants of low imputation quality, contributed to retaining information. Our supporting statistical tests enabled us to detect statistically significant differences in genetic covariance and its standard error while considering the varying number of shared genetic variants across bivariate trait pairs.

**Conclusion:** LDSC++ was confirmed to produce less biassed estimates of genetic covariance and its variability in our GLAD+ sample compared to standard LD score regression, using GCTA-REML as reference. This performance was supported by results from external trait GWASs of varying character, also implying an important performance of our extended weighting schemes. Our proposed extensions to LD score regression, among which genome-wide parameters are constructed as aggregates of heterogeneous local parameters, may prove important for large-scale multivariate studies such as genomic structural equation models or local genetic covariance analyses.

## 1 INTRODUCTION

Genome-wide association study (GWAS) summary statistics are readily available to researchers worldwide compared to individual level genotyped or sequenced data, which typically require considerable cost and time to access due to data sharing restrictions^1,2^. This prevents the use of optimal methods that use individual level genotype data to estimate components of variance, such as GCTA-GREML^3,4^, BOLT-REML^5,6^ and REML in LDAK^7^. Popular alternatives^8^ are Linkage Disequilibrium (LD) score regression^9^ (LDSR) and SumHer^10^, which instead use GWAS summary statistics and an LD reference panel to obtain common genetic variant covariance (covG) and heritability (h^2^), which are standardised to estimate genetic correlations^11^ (r_g_). LDSR was later re-implemented for calculation of large-scale bivariate trait pair analyses to obtain covG matrices and variance estimates of the elements of these for genomic structural equation modelling in Genomic SEM^12^.

However, available implementations of LDSR do not: 1.) explicitly account for varying numbers of genetic variants between association statistics from contributing GWASs, 2.) consider local association and LD patterns in GWASs, or 3.) adjust for imputation quality differences. In bivariate LDSR, genetic variant overlap between two traits and the LD score panel is usually drastically smaller compared to the regressions performed on single traits (supplementary tab. S6), due to differing patterns of missingness across datasets. This limits the power to carry out many downstream multivariate GWAS analyses, such as in Genomic SEM^12^, and related methods such as mtCOJO^13,4^, MTAG^14^, and METAL^15^. These methods rely on the covG estimates from LDSR, and/or use the LDSR intercept to adjust associations (genomic control) for spurious associations due to confounding from population stratification or sample overlap. In comparison, the variance-component REML method in LDAK^7^ is both assuming a variable contribution to heritability across the genome and includes an information score weighting^16^ to differentially treat variants of varying genotyping or imputation quality. It may be of interest to bring this functionality over to an LDSR setting to be applicable to summary level data rather than individual level data. Standard LDSR further assumes there to exist a heteroskedastic relationship between genome-wide covG and its standard error, mainly due to the expected larger variability of variants in high LD regions. Despite a weighting correction to account for genome-wide covG heteroskedasticity, it also served as justification^9^ for employing resampling to estimate standard errors that are robust to bias from heteroskedasticity. It may therefore be of interest to investigate whether this assumption can be relaxed when estimating covG standard errors, to allow for sampling across the genome.

Here we present an enhanced version of LDSR, LDSC++, extended to improve on 1.) the handling of varying numbers of shared genetic variants across trait pairs and reference panels, 2.) the estimation of genetic covariance and its standard error, and 3.) the handling of imputation quality of imputed genotypes or genetic associations. Finally, through our extensions we aim to allow for tractable statistical tests of covG, its standard error, or differences between them, also accounting for high correlations between compared variables. We validate the performance of our extensions to LDSR, first using simulations, second on real-world data in the form of GWASs performed on selected measurements in the Genetic Links to Anxiety and Depression study and the United Kingdom National Institute of Health and Social Care Research BioResource, and third on a selection of ten GWAS summary statistics representing a wide variety of traits. We hypothesise the accuracy of estimating both covG and its standard error may be improved and the average measured variability of covG estimates across a sample of multiple diverse traits will be reduced when applying our extensions in LDSC++ individually or in combination, as compared to standard LDSR.

## 2 MATERIALS AND METHODS

To enable more precise covG estimation from GWAS summary statistics, we extend the LDSR method based on the multivariate implementation included in Genomic SEM. We have named this extension LDSC++ (supplementary info. 1.8). The extensions to LDSR are:

❖ **Extension 1 - Variable block-count:** Allowing the number of blocks to vary when estimating covG and covG standard error, in contrast to a fixed number of blocks.
❖ **Extension 2 - Extended block definitions to allow for variable block-count:** Block definitions to be used while allowing the number of blocks to vary with Extension 1. These are based on either a set number of variants or a recombination distance in cM.
❖ **Extension 3 - Variable block-count sampling:** A novel approach to sample across genome blocks to estimate genome-wide covG and covG variance, rather than performing block jackknife resampling.
❖ **Extension 4 - Extended weighting scheme for imputation quality:** An addition to the standard LDSR weighting scheme to also control for imputation quality.
❖ **xtension 5 - Adjusted weighting scheme to correct for the correlation between LD score and association statistic:** We adjusted the standard weighting scheme in LDSR to correct for correlation between the regression variables, to allow for LD scores smaller than one while preventing influences by extreme LD (close to zero) values due to noise.

LDSC++ alongside standard LDSR (emulated in our LDSC++ code) were validated in both real-world data and simulated traits, using REML covG estimates for reference.

An individual level sample from cohorts part of the National Institute of Health Research (NIHR) BioResource was used for assessing genetic associations with phenotypes and for simulation. This selection included participants of European ancestry from the Genetic Links to Anxiety and Depression study (GLAD)^17^, the United Kingdom Eating Disorders Genetics Initiative (EDGI)^18,19^, the NIHR BioResource Inflammatory Bowel Disease BioResource panel and the NIHR BioResource General Population panel, which together form a collection of cohorts colloquially referred to as GLAD+ (N=36,746).

We selected the traits height, weight, neuroticism measured by the Eysenck Personality Questionnaire-Revised Short form (EPQR-S)^20^, and negative affect measured by the PID-5 Brief Form^21,22^, in the GLAD+ cohort. Additionally, we simulated two phenotypes (further referred to as SIM1 and SIM2 respectively) created to have 20% heritability and a genetic correlation of 0.4 with each other, in four pairs with fractions of 100%, 75%, 50%, and 25% shared quantitative trait loci (QTL) respectively. A simulation was performed in GCTA^4^ for each phenotype and fraction combination, using 30,000 QTL and GLAD+ as an individual level sample.

We used GCTA-GREML^3^ implemented in GCTA^4^ to estimate variance component covG for our individual level GLAD+ samples. Association tests were performed in GCTA fastGWA^23^. The covariates included in the association analysis were: genotyping array type/version, tissue source, genotyping batch, and the first seven principal component scores of participant genotypes as computed by the genetic principal component eigenvalues of the 1000 Genome Project^24^ Phase 3 (1kG3) reference panel projected on the genetic variants of GLAD+. Additionally, ten external GWAS summary statistics covering a range of traits of various statistical power, h^2^, and genome coverage in terms of the number of variants included were selected for validating methods (Tab. 1). All summary statistics were documented to be based on samples with predominantly European ancestry.

**Table 1.** Selected external GWAS summary statistics. A selection of ten diverse publicly available GWAS summary statistics. The table describes the dataset reference, the number of cases or total number of participants, and for binary traits; the number of controls, the sample prevalence estimate, and the population prevalence estimate used throughout the study. ‘INFO range’ describes the range of the provided imputation quality information score.

| Trait | Reference | Year | N case | N control | Sample prev. | Population prev. | INFO range |
| --- | --- | --- | --- | --- | --- | --- | --- |
| Anxiety disorder (ANXD) | Purves et al.<sup>35</sup> | 2019 | 25,453 | 58,113 | 0.305 | 0.16<sup>36</sup> | 0.8-1.0 |
| Bipolar disorder (BIPD) | Stahl et al.<sup>37</sup> | 2019 | 20,352 | 31,358 | 0.394 | 0.025<sup>38</sup> | 0.1-1.6 |
| BMI (BMI) | Pulit et al.<sup>39</sup> | 2019 | 806,834 |  |  |  | 0.3-1.0 |
| MDD (MDD) | Wray et al.<sup>40</sup> | 2018 | 16,823 | 25,632 | 0.396 | 0.146<sup>41</sup> | 0.6-1.9 |
| Broad depression (MDDb) | Howard et al.<sup>42</sup> | 2019 | 170,756 | 329,443 | 0.341 | 0.302<sup>42</sup> | - |
| MDD symptoms (MDDS) | Wray et al.<sup>40</sup> | 2018 | 14,260 | 15,480 | 0.479 | 0.146<sup>41</sup> | 0.9-1.0 |
| Educational attainment (EDUA) | Lee et al.<sup>43</sup> | 2018 | 766,345 |  |  |  | - |
| Insomnia (INSO) | Jansen et al.<sup>44</sup> | 2019 | 109,402 | 277,131 | 0.283 | 0.69<sup>45</sup> | 0.9-1.0 |
| Neuroticism (NEUR) | Nagel et al.<sup>46</sup> | 2018 | 449,484 |  |  |  | 0.9-1.0 |
| Schizophrenia (SCHI) | Pardiñas et al.<sup>47</sup> | 2018 | 40,675 | 64,643 | 0.386 | 0.0072<sup>48</sup> | - |

To match GCTA-GREML, we used GCTA^4,25^ to generate the GLAD+ LD scores, using a window of 1 Mb and a prior filter on minor allele frequency (MAF) > 0.01. For our external GWAS summary statistics, we instead created LD scores based on the European ancestry subset of the 1kG3^26^, not including Finnish ancestry. For this second LD score library, we used a 1 cM window and 250 blocks setting in LDSC^9,27^.

GWAS summary statistics were first harmonised using our *Supermunge* routine part of the *shru*^28^ package for the R language [https://github.com/tnggroup/tng_fork_shru_jz2024], based on the original LDSC munge routine^9^, the Genomic SEM routines for munging and preparation for latent factor GWAS^12^, and the MungeSumstats routine^29^ (supplementary info. 1.4.4).

To account for heteroskedastic relationships for comparisons of covG inferential uncertainty (sampling variance and thus standard errors) across traits of varying heritabilities, we applied the coefficient of variation (CV) of covG rather than comparing direct estimates (supplementary info. 1.5.1).

We formulated simple parametric statistical tests specifically accounting for the novel varying number of blocks in the LDSR sampling and resampling methods and non-independent variables (supplementary info. 1.10), supported by paired samples Wilcoxon signed-rank tests^30^ (supplementary info. 1.10.5). Test results were corrected for multiple comparisons using the Benjamini & Hochberg^31^ method (q-values, also called false discovery rate; FDR), additionally and unconventionally taking the effective number of comparisons from correlated variables^32–34^ into account (supplementary info. 1.10.6).

## 3 RESULTS

We estimated REML h^2^ and r_g_ of our four selected GLAD+ traits and their bivariate combinations. The REML h^2^ of the simulated traits SIM1 and SIM2 were estimated in the range from 13% to 15%, and the REML r_g_ between traits in a pair to 0.43, 0.24, 0.19, and 0.2 for the QTL sharing fractions 100%, 75%, 50%, and 25% respectively. Deviations of LDSR method results from GCTA-REML estimates were obtained, run on both the selected GLAD+ traits (Tab. 2, supplementary tab. S7A, S7B) and the simulated traits (supplementary tab. S8A, S8B). Most deviations between covG and covG variance estimates by our LDSR method extensions compared to standard LDSR were seen to be statistically significant, supporting our extensions to yield empirically distinct results. As results were quite different comparing results from diagonal and off-diagonal elements of covG matrices, we decided to stratify them accordingly.

**Table 2.** LDSR estimate deviation from a REML ideal measured over four GLAD+ traits. The table shows deviations of LD score regression (LDSR) method estimates of genetic covariance (covG) and covG SE (converted to coefficient of variation, CV) from the chosen ideal; estimates from GCTA-GREML using individual level data. We used the LDSC++ implementation of LDSR applied to our selection of four real-world traits from the GLAD+ sample: height, weight, neuroticism measured by the Eysenck Personality Questionnaire-Revised Short form (EPQR-S), and negative affect measured by the PID-5 Brief Form. Deviation was measured as average relative difference (Avg. Δ) and root-mean-square error (RMSE). LDSR method combinations were defined by their configuration in regard to block definition, (re)sampling method, alternative correlation correction weighting, and imputation quality weighting. Estimates are stratified in diagonal (heritability) and off-diagonal (bivariate genetic covariance) estimates from a genetic covariance matrix. Only significant covG values (q<0.05) were considered.

| Block definition | (Re)sampling method | Alternative correlation correction weighting | Imputation quality weighting | covG |  |  |  | covG CV |  |  |  |
| --- | --- | --- | --- | --- | --- | --- | --- | --- | --- | --- | --- |
|  |  |  |  | Diagonal |  | Off-diagonal |  | Diagonal |  | Off-diagonal |  |
| | | | | Avg. $\Delta$ (%) | RMSE | Avg. $\Delta$ (%) | RMSE | Avg. $\Delta$ (%) | RMSE | Avg. $\Delta$ (%) | RMSE |
| Fixed 200 blocks | Block jackknife resampling | ☐ | ☐ | -32.7 | 0.0754 | -61.0 | 0.0994 | 7.6 | 0.0756 | 118.0 | 0.1283 |
| Number-of-variants blocks | Block jackknife resampling | ☐ | ☐ | -32.7 | 0.0754 | -61.0 | 0.0994 | 7.9 | 0.0788 | 120.0 | 0.1300 |
|  |  | ☒ | ☐ | -33.0 | 0.0759 | -61.3 | 0.0998 | 8.7 | 0.0769 | 122.4 | 0.1327 |
|  |  | ☐ | ☒ | -33.0 | 0.0756 | -61.3 | 0.0998 | 6.4 | 0.0769 | 116.5 | 0.1231 |
|  |  | ☒ | ☒ | -33.2 | 0.0760 | -61.5 | 0.1002 | 7.3 | 0.0750 | 118.8 | 0.1256 |
|  | Variable block-count sampling | ☐ | ☐ | -21.7 | 0.0542 | -50.1 | 0.0836 | -5.8 | 0.1182 | 79.4 | 0.0783 |
|  |  | ☒ | ☐ | -22.0 | 0.0549 | -50.4 | 0.0841 | -5.6 | 0.1173 | 80.5 | 0.0795 |
|  |  | ☐ | ☒ | -21.0 | 0.0523 | -52.3 | 0.0861 | -6.7 | 0.1185 | 81.4 | 0.0807 |
|  |  | ☒ | ☒ | -21.2 | 0.0529 | -52.6 | 0.0867 | -6.5 | 0.1175 | 82.8 | 0.0821 |
| Recombination distance blocks | Block jackknife resampling | ☐ | ☐ | -31.7 | 0.0722 | -59.8 | 0.0972 | 5.1 | 0.0727 | 101.8 | 0.1155 |
|  |  | ☒ | ☐ | -31.9 | 0.0726 | -60.0 | 0.0976 | 5.9 | 0.0707 | 103.6 | 0.1176 |
|  |  | ☐ | ☒ | -31.9 | 0.0721 | -60.0 | 0.0975 | 5.7 | 0.0703 | 103.1 | 0.1177 |
|  |  | ☒ | ☒ | -32.1 | 0.0726 | -60.2 | 0.0979 | 6.5 | 0.0683 | 105.0 | 0.1198 |
|  | Variable block-count sampling | ☐ | ☐ | -11.1 | 0.0475 | -38.5 | 0.0638 | -1.6 | 0.1132 | 59.7 | 0.0925 |
|  |  | ☒ | ☐ | -11.4 | 0.0480 | -38.5 | 0.0643 | -1.1 | 0.1127 | 60.9 | 0.0944 |
|  |  | ☐ | ☒ | -11.2 | 0.0482 | -46.4 | 0.0784 | -1.4 | 0.1134 | 99.0 | 0.0962 |
|  |  | ☒ | ☒ | -11.5 | 0.0487 | -39.3 | 0.0647 | -0.8 | 0.1129 | 62.9 | 0.0910 |

Only considering h^2^ estimates (diagonal elements in the covG or covG SE matrix), the top performing LDSR method combination was variable block-count sampling using the recombination distance block-definition, which yielded average relative h^2^ differences as small as -11% and average relative h^2^ CV differences of -1% compared to GCTA-GREML in GLAD+. The same pattern was seen for the simulated traits only using the 100% QTL sharing fraction, while for lesser sharing fractions the performance among methods was mixed with the standard LDSR block jackknife or LDSC++ variable block-count sampling with recombination distance blocks yielding the on average most accurate and less biassed estimates. In the GLAD+ data, all LDSR methods under-estimated diagonal covG with about -30% for block jackknife resampling and - 10% to -20% for variable block-count sampling. Block jackknife resampling was also consistently seen to over-estimate diagonal variability estimates from 5% to 9%, while variable block-count sampling seemed to under-estimate diagonal variability estimates with about -1% to -7%. The influence of our weighting extensions were seen to only have tiny effects on the differences in the preferred direction for the GLAD+ traits, primarily detectable for h^2^ estimates and when using block jackknife resampling and imputation quality weighting.

Off-diagonal deviations were markedly larger by about a factor of two to four for average relative covG estimates and a factor of ten to sixty for average relative covG CV estimates, compared to the corresponding diagonal matrix element results for the GLAD+ traits. A similar but attenuated pattern was seen for the simulated traits. As for the diagonal elements, the LDSR methods under-estimated off-diagonal covG estimates with about -60% for block jackknife resampling and -40% to -50% for variable block-count sampling in the GLAD+ traits. Off-diagonal covG CVs were consistently and grossly over-estimated by all LDSR methods. For block jackknife resampling, the recombination distance block definition brought the off-diagonal covG CV difference to about 100% compared to 120% when using number-of-variants defined blocks. Variable block count sampling produced off-diagonal covG CV estimates with deviations from about 60% with the recombination distance block definition to 80% with the number-of-variants block definition.

We applied the different LDSR method combinations in our selection of ten external trait GWASs (Tab. 1) for which samples we did not have individual level data (supplementary tab. S9A, S9B). Default block-size settings resulted in block-counts in the range of 249 - 405 for the number-of-variants block definition and 338 - 364 for the recombination distance block definition, without INFO filter. Estimates from method combinations using our extensions were first assessed independently in reference to suitable baselines (supplementary tab. S9A). Compared to standard LDSR, applying our block definitions and sampling extensions yielded 4% to 8% larger h^2^ estimates, 3% smaller h^2^ CV estimates using number-of-variants blocks, but 7% larger h^2^ CV estimates using recombination distance blocks. The weighting extensions made a relatively small contribution to average differences in estimates compared to the block and sampling extensions, but specific traits showed statistically significant results. Applying imputation score weighting resulted in an approximate gain of 4% to 5% (q<0.05) in BIPD h^2^ and a 1% to 7% (q<0.02) reduction in BIPD bivariate covG CV with specific traits across block and sampling methods. Similarly, applying alternative correlation correction weighting yielded an approximate gain of 3% to 4% (q<0.05) in BMI h^2^ and multiple significant bivariate BMI covG CV reductions in the range of 1% to 10% (q<0.05). Applying all extensions in concert and comparing to standard LDSR yielded 5% to 8% larger h^2^ estimates, 4% smaller h^2^ CV estimates using number-of-variants blocks, but 6% larger h^2^ CV estimates using recombination distance blocks.

## 4 DISCUSSION

We have adapted the LDSR routine from Genomic SEM into a modified implementation called LDSC++, which performs LDSR with extensions specifically aimed at improving covG estimation in large-scale multivariate analyses with varying genomic overlaps among trait GWAS dataset and reference panels. For this purpose, we created a variable block-count sampling routine in LDSC++ aimed to be applied with a variable number of blocks to discriminate between situations with a varying number of participating genetic variants, which further makes it possible to use this information in statistical tests. Variable block-count sampling performs segmented (also called piecewise) regression using one of our two block definitions, which putatively makes the aggregate genome-wide parameter estimates (averages of segmented regression parameters) robust to heteroskedasticity and outliers. The most obvious and important assumption for both the variable block-count sampling method and the statistical tests is that the estimated genome-wide covG can be assumed to have the same variance across the genome, or rather can be estimated as an aggregate of piecewise estimates. While resampling techniques^49,50^ traditionally have been used to obtain robust covG standard errors in LDSR, a sufficiently better model from the segmented regression is in our project theorised to make robust standard errors superfluous^51^. LDSC++ further takes advantage of imputation quality information to weight the LDSR, instead of relying on filters which otherwise reduce the overall information content in the analysis. Variable block-count sampling may be interpreted as applying a slightly different h^2^ model, allowing for a different contribution to h^2^ across the genome similar to the LDAK h^2^ model^7,16,52^, in contrast to the standard LDSR heritability model^9,16,52^. We additionally proposed statistical tests in which the varying number of blocks from our extensions can be applied to translate statistical power from a large number of genetic variants in a GWAS into statistical power for LDSR covG and covG variance, and thus making these tests more sensitive and differentiating in collections of bivariate trait pairs proportional to the variability in genetic variant overlap.

Our LDSR extensions were shown to yield more accurate results as validated by GCTA-GREML than standard LDSR settings, both when using each extension independently and when applied in concert, yielding average relative deviations from about -10% to -20% for h^2^ and -1% to -6% for h^2^ CV compared to the corresponding deviations of -30% and 8% respectively for standard LDSR. This performance was seen in estimates made in the individual level GLAD+ sample and replicated to a lesser extent in simulated data using GCTA-GREML. The performance increase when using the combined extensions could be attributed to allowing the number of blocks to vary and using our new block definitions or using our variable block-count sampling, and only a small fraction could be attributed to the weighting extensions in this setting. Applying our extensions to a set of ten trait GWASs external to our individual level sample, supported the above validations through convergent results primarily for h^2^ estimates rather than off-diagonal covG estimates and when using number-of-variants block definitions. In this diverse selection of GWASs of varying power and coverage our combined LDSC++ extensions yielded on average up to 8% larger and 4% less variable h^2^ estimates compared to standard LDSR. Larger h^2^ estimates were convergent with the previous individual level data validation validations, but only the number-of-variants block definition were seen to produce smaller average h^2^ estimates compared to standard LDSR, while using the recombination distance block definition instead yielded larger average h^2^ estimates - contrary to the previous validation results. This may indicate a sensitivity of the recombination distance block definition to missingness patterns or other factors that were not represented by our four GLAD+ trait GWASs. The external GWASs also allowed us to explore other imputation score ranges and highly polygenic trait GWASs of high power, which implicated putative improved estimates when applying the imputation score weighting to the BIPD GWAS and when applying the alternate correlation correction weighting to the BMI GWAS. Our proposed explanation for these results is that the BIPD GWAS has a wide distribution of imputation quality INFO scores in contrast to other included datasets which the corresponding weighting extension would be able to leverage, and the BMI GWAS is relatively well-powered and represents a highly polygenic trait for which the alternative correlation correction weighting may be able to correct bias due to tiny LD scores to a higher degree.

The major strength in our simulations came from using a relatively large sample size of 36,746 individuals from the GLAD+ cohort, in contrast to the Bulik-Sullivan study which simulated associations based on a sample of 2,062 individuals. We additionally simulated whole genome associations, where the Bulik-Sullivan study only simulated chromosome 2. Unfortunately, we did not manage to perform sufficiently many repetitions in our simulations for them to appear trustworthy, with an effective number of GCTA simulations of 2.5 for any randomly sampled QTL compared to 100 repetitions done in a previous study by Bulik-Sullivan^53^. Our effort to investigate the influence of effect QTL overlap on REML and LDSR estimates was interesting, but proved practically challenging enough to be one of the reasons for why we did not manage to repeat the simulations to a higher degree.

The discrepancy between validation results from diagonal and off-diagonal covG elements was identified as a cause for concern about the LDSR method in general. Because of this we were inclined to trust off-diagonal validation results less. Off-diagonal covG estimates from bivariate LDSR are however the foundation for genetic correlation analyses, and also provide the majority and arguably the more important information for analyses performed on covG such as Genomic SEM, underscoring the importance of accurate off-diagonal covG estimation and the need to critically investigate bivariate LDSR further.

Our results provide support for the accuracy of statistical models where genome-wide h^2^ and covG parameters are constructed as aggregates of heterogeneous local parameters. The recombination distance block definition was part of the method combination displaying the strongest performance in our individual level real-world and simulated GCTA-GREML validations. In the external GWASs, the recombination distance block definition also recovered some of the largest covG estimates on average compared to the number-of-variants block definition and standard LDSR. We would suggest that these properties are promising and may prompt further research into block definitions for LDSR that are consistent across traits and bivariate trait pairs in the way our recombination distance definition was made or how local genetic correlation methods such as LAVA^54^ define consistent genomic blocks. The arbitrary definition of blocks is a general limitation of LDSR. Creating more optimised block definitions is indeed an active research area^55,56^. Reconciling the recombination based block definition approach with the utility of the number-of-variants block definition may be important to advance LDSR and similar methods further.

## 6 ACKNOWLEDGEMENTS

This study/research is funded by the National Institute for Health and Care Research (NIHR) Maudsley Biomedical Research Centre (BRC). The views expressed are those of the author(s) and not necessarily those of the NIHR or the Department of Health and Social Care.

Christopher Hübel acknowledges funding by Lundbeckfonden (R276-2018-4581).

Alexandra Gillett is supported by the Medical Research Council (MR/X009815/1).

The authors acknowledge use of the research computing facility at King’s College London, King’s Computational Research, Engineering and Technology Environment (CREATE) (https://doi.org/10.18742/rnvf-m076)^57^.

## 7 DATA AVAILABILITY

All data produced in the present study are available upon reasonable request to the authors. While some data is contained in the manuscript, an attempt has been made to additionally present results in table format in the accompanying supplementary tables. GWAS summary statistics of our traits used for validation are available upon reasonable request. LD scores based on the GLAD+ sample are available upon reasonable request. The analysis code will be made available at https://github.com/tnggroup/jz-phd-c2c3-2024. The software package ‘shru’ will be made available on https://github.com/johanzvrskovec/shru.

## 8 ETHICS APPROVAL STATEMENT

The London—Fulham Research Ethics Committee approved the GLAD Study on 21st August 2018 (REC reference: 18/LO/1218). The London—Fulham Research Ethics Committee approved the EDGI UK Study on 29th July 2019 (REC reference: 19/LO/1254). The East of England— Cambridge Central Committee approved the NIHR BioResource as a Research Tissue Bank (REC reference: 17/EE/0025). The South West—Central Bristol Research Ethics Committee approved the COVID-19 Psychiatry and Neurological Genetics study on 27th April 2020 (REC reference: 20/SW/0078).

