## Supplementary information for "LDSC++: Improving linkage disequilibrium score regression estimation of heritability and genetic correlation for multivariate GWAS analysis"

### 1 Supplementary methods

#### 1.1 General

Please note that since we have based our comparisons on the implementation of LD score regression (LDSR) in Genomic SEM^1^, any reference to standard LDSR, or just LDSR, is to the Genomic SEM implementation if not otherwise specified.

#### 1.2 Software and computation infrastructure

Most analysis was performed in R^2^ with major version 4. For computationally intensive or memory demanding operations we relied on King's Computational Research, Engineering and Technology Environment (CREATE)^3^.

#### 1.3 Individual level sample

We used an individual level sample from cohorts part of the National Institute of Health Research (NIHR) BioResource. This selection included participants from the Genetic Links to Anxiety and Depression study (GLAD)^4^, the United Kingdom Eating Disorders Genetics Initiative (EDGI)^5,6^, the NIHR BioResource Inflammatory Bowel Disease BioResource panel and the NIHR BioResource General Population panel, which together form a collection of cohorts colloquially referred to as GLAD+. At the time of our analyses, the collective GLAD+ cohort included 36,746 genotyped and quality controlled participants. The sample was confirmed to be of predominantly European ancestry as defined by genetic principal component analysis, and further analysis was restricted to included only study participants of European ancestry defined in this way. Self-report of measurements through online questionnaires were predominantly used for phenotypic data collection in GLAD+.

#### 1.4 GWAS summary statistics

##### 1.4.1 Simulation of genetic associations from individual level data

We simulated two phenotypes (further referred to as SIM1 and SIM2 respectively) defined by their effect sizes on a selection of genetic variants from our reference panels (see Reference and LD score panels below). The simulated phenotypes were created to have 20% heritability and 40% genetic correlation with each other, in four pairs with fractions of 100%, 75%, 50%, and 25% shared quantitative trait loci (QTL) respectively. A simulation was performed in GCTA^7^ for each phenotype and fraction combination, using 30,000 QTL and a specified 20% heritability. Association tests were performed in GCTA fastGWA^8^. The GLAD+ cohort was used as an individual level sample for the simulated GWASs.

##### 1.4.2 Genome-wide association analysis of traits in GLAD+

We performed simple association analyses of a selection of well-powered traits measured in the GLAD+ cohort, to serve as a basis for comparisons with REML methods applied on the individual level data for the same variables. We selected the traits height (in m), weight (in kg), neuroticism measured by the Eysenck Personality Questionnaire-Revised Short form (EPQR-S)^9^, and negative affect measured by the PID-5 Brief Form^10,11^. The covariates included in the association analysis were: genotyping array type/version, tissue source, genotyping batch, and the first seven principal component scores of participant genotypes as computed by the genetic principal component eigenvalues of the 1000 Genome Project^12^ Phase 3 (1kG3) reference panel projected on the genetic variants of GLAD+. These association analyses were solely performed for comparison reasons to evaluate the performance of LDSC++ and should not be preferred over results from association analyses performed on the same sample that have taken measures to avoid selection bias from for example sex or diagnosis status.

##### 1.4.3 Selection of real world GWAS summary statistics

We selected 10 GWASs covering a range of psychiatric, behavioural and anthropometric traits of various statistical power, h^2^, and genome coverage in terms of the number of variants included (Tab. 1, supplementary tab. S4). All summary statistics were documented to be based on samples with predominantly European ancestry.

**Table 1. Selected GWAS summary statistics**

The table describes the dataset reference, the number of cases or total number of participants, and for binary traits; the number of controls, the sample prevalence estimate, and the population prevalence estimate used throughout the study. ‘INFO range’ describes the range of the provided imputation quality information score.

| Trait | Reference | Year | N case | N control | Sample prev. | Population prev. | INFO range |
| --- | --- | --- | --- | --- | --- | --- | --- |
| Anxiety disorder (ANXD) | **Purves et al.**^13^ | **2019** | **25,453** | **58,113** | **0.305** | **0.16**^14^ | **0.8-1.0** |
| Bipolar disorder (BIPD) | **Stahl et al.**^15^ | **2019** | **20,352** | **31,358** | **0.394** | **0.025**^16^ | **0.1-1.6** |
| BMI (BMI) | **Pulit et al.**^17^ | **2019** | **806,834** |  |  |  | **0.3-1.0** |
| MDD (MDD) | **Wray et al.**^18^ | **2018** | **16,823** | **25,632** | **0.396** | **0.146**^19^ | **0.6-1.9** |
| Broad depression (MDDB) | **Howard et al.**^20^ | **2019** | **170,756** | **329,443** | **0.341** | **0.302**^20^ | **-** |
| MDD symptoms (MDDS) | **Wray et al.**^18^ | **2018** | **14,260** | **15,480** | **0.479** | **0.146**^19^ | **0.9-1.0** |
| Educational attainment (EDUA) | **Lee et al.**^21^ | **2018** | **766,345** |  |  |  | **-** |
| Insomnia (INSO) | **Jansen et al.**^22^ | **2019** | **109,402** | **277,131** | **0.283** | **0.69**^23^ | **0.9-1.0** |
| Neuroticism (NEUR) | **Nagel et al.**^24^ | **2018** | **449,484** |  |  |  | **0.9-1.0** |
| Schizophrenia (SCHI) | **Pardiñas et al.**^25^ | **2018** | **40,675** | **64,643** | **0.386** | **0.0072**^26^ | **-** |

##### 1.4.4 Preparation of GWAS summary statistics

We developed a quality control and harmonisation routine; *Supermunge* which is part of the *shru*^27^ package for the R language [<https://github.com/tnggroup/tng_fork_shru_jz2024> ], based on the original LDSC munge routine^28^, the Genomic SEM routines for munge and preparation for latent factor GWAS^1^, and the MungeSumstats routine^29^. Supermunge was created to resolve issues with variant naming and/or position duplications, to interpret allele order and sign direction while treating any effect direction as equally possible, and to infer missing data from a provided variant reference, in addition to interpret a large number of existing tab separated value file formats for GWAS summary statistics. Supermunge resolves duplicate occurrences of a variant name/ID by saving the variant with the position closest to the position as specified by the reference, or otherwise with the highest MAF. Duplicates are excluded after joining with the LD scores to avoid duplicated variants due to multiple occurrences in either of the GWAS summary statistics, reference panel harmonisation (‘munge’) variant list, or the LD score panel. Supermunge was first implemented in R version 4.2.0 as part of a publicly available R package containing various R utilities for statistical genetics and structural equation modelling, and makes extensive use of the data.table^30^ R package to achieve performance in terms of processing speed. The standardised variable names used in Supermunge are described in the supplementary tables (S1) together with an overview of the implemented processing steps (S2). Users of Supermunge may use tab separated value files for reference variant lists and GWAS summary statistics that are either gzipped or in plain text which use column names as described in the supplement. Supermunge additionally has an extensive column interpreter that will attempt to convert any input file format to the column standard and is able to read common formats such as the ‘Daner’ or summary VCF type files that are commonly used by the Psychiatric Genomics Consortium^31^.

GWAS summary statistics were first harmonised using Supermunge with the features described above, highlighting the functionality to resolve duplicate variant occurrences. Effective sample size^32^ was used for binary traits in LDSR and conversions from observed scale to liability scale covG were performed using published lifetime population prevalences (Tab. 1). Strict ordering of the variants in the GWAS summary statistics was additionally enforced according to chromosome number, base-pair position, and variant name (rs-id usually), which was expected to harmonise LDSR across traits using either a fixed number of blocks or blocks defined by number of variants. Variants in GWAS summary statistics with MAF <0.01 were excluded before LDSR. Similarly, variants with INFO < 0.6 were excluded when not applying our imputation quality weighting scheme, if not otherwise specified.

#### 1.5 Relations used for comparison and validation

##### 1.5.1 Coefficient of variation

We measured the correlation between LDSR covG and its standard errors in our selection of real world trait GWASs to 0.66 (p=4.6e-8). Based on this we further made the assumption that the data generating process underpinning covG generates a heteroskedastic relationship between covG and its standard error, where this process generating covG of larger magnitudes would also generate variance (and thus standard errors) of larger magnitudes. Any within-genome heteroskedasticity of covG can be assumed to at least extend, and at most be exaggerated, across traits. A standard way to compare variability across measurements expected to have a size dependent type of heteroskedasticity is to use the coefficient of variation^33^ (CV), which we used for all comparisons between covG variability (variance or standard errors) estimated from LDSR. A CV is generally defined as the ratio of a standard deviation $\sigma$ to the absolute mean of its distribution $\mu$ as $CV=\frac{\sigma}{\left| \mu\right|}$ .

We further capped the covG used in the denominator to a minimum of 0.05 of the corresponding genetic correlation to avoid the known problem of the CV becoming severely inflated as the covariance denominator approaches 0, sometimes due to noise. For a genetic covariance from any of the above LDSR estimators $\hat{\rho}_{g,any}$, we constructed the corresponding CV with a capped covG denominator as:

$\Sigma_{\hat{\rho}_{g,any}}=\frac{S_{\hat{\rho}_{g,any}}^{2}}{\lambda_{g}}$	(Formula 5.1)

To avoid problems with infinitely small covG estimates, the standard error is standardised using a capped representation of covG as:

$\lambda_{g}=\left\{ \frac{\left| \hat{\rho}_{g,any} \right|}{\sqrt{\hat{\rho}_{g1diag}}\sqrt{\hat{\rho}_{g2diag}}}\geq C_{lim} : \left| \hat{\rho}_{g,any} \right| ,\frac{\left| \hat{\rho}_{g,any} \right|}{\sqrt{\hat{\rho}_{g1diag}}\sqrt{\hat{\rho}_{g2diag}}}<C_{lim} :C_{lim}\sqrt{\hat{\rho}_{g1diag}}\sqrt{\hat{\rho}_{g2diag}} \right\}$	(Formula 5.2)

where $\hat{\rho}_{g1diag}$ and $\hat{\rho}_{g2diag}$ respectively are the corresponding covG of trait 1 and 2 with themselves (i.e. their heritabilities), and $C_{lim}$ is a lower cap on the scale of genetic correlation. We have used a default value of 0.05 for $C_{lim}$.

##### 1.5.2 Root mean square error

The root mean square error (RMSE), also known as the root mean square deviation (RMSD), is a common measure of deviance from an ideal. We used RMSE as a measure of deviance when validating our methods. RMSE is defined as:

$RMSE=\sqrt{\frac{\sum_{n}^{N} \left( x_{n}-y_{n} \right)^{2}}{N}}$	(Formula 5.3)

Here $x_{n}$ is a measurement from a set of values we want to validate against another set of paired values $y_{n}$, which are considered to be the reference (in our study the reference is ‘The Truth’ estimates of covG, h^2^, and their standard errors, obtained from GCTA-GREML). The size of the paired value set is $N$.

#### 1.6 Reference and LD score panels

To match the GCTA-GREML method (see Variance component covG estimation) which uses in-sample LD, we used the GLAD+ cohort individual level data as genetic variant reference, and to generate in-sample LD scores for LDSR of our simulated traits. To further match GCTA-GREML, we used GCTA^7,34^ to generate the GLAD+ LD scores, using a window of 1 Mb and a prior filter on minor allele frequency (MAF) > 0.01. For our selected real world data, we instead created LD scores based on the European ancestry subset of the 1000 Genomes Project^35^ phase 3 reference panel (1kG3), not including Finnish ancestry. For this second LD score library, we used a 1 cM window and 250 blocks setting in LDSC^28,36^. The 1kG3 panel was filtered to variants with MAF > 0.001 before generating LD scores (supplementary tab. S5B).

#### 1.7 Variance component covG estimation

We used GCTA-GREML^37^ implemented in GCTA^7^ to estimate variance component covG of common genetic variants. GCTA-GREML relies on inferring covG from a genetic relatedness matrix (GRM) and further corrects for the in-sample LD^38^. Similar to LDSR, GCTA-GREML assumes a uniform heritability model where the expected heritability is not proportional to neither the minor allele frequency (MAF) nor the level of LD. Due to REML estimates being based on individual level data and thus considered to be more accurate, they were used in later validations of our LDSR extensions, as references to represent ideal target estimates (‘The Truth’).

#### 1.8 LDSC++: LD score regression adapted for multivariate GWAS and varying variant-count

##### 1.8.1 The LDSR formula

In LDSR^28,39^ the Z test statistics of a GWAS are regressed on the LD scores of the corresponding markers. It is used to estimate the genetic covariance (covG, also denoted $\rho_{g}$) between the two traits according to the following linear relationships:

$Cov\left( Z_{1j},Z_{2j} \right)=E\left[ Z_{1j}Z_{2j}|l_{j} \right]= \frac{\sqrt{N_{1}N_{2}}}{M_{LDSR}}\rho_{g}l_{j}+1+ \zeta$	(Formula 8.1)

Here, for a variant $j$,

$Z_{1j}Z_{2j}$ are the association weighted test Z statistics for each of the two traits with sample sizes $N_{1j}N_{2j}$,

$l_{j}$ is the weighted LD score,

$M_{LDSR}$ is the number of markers that were used to contribute to the LD scores (which may further have been filtered to MAF > 0.05),

$\rho_{g}$ is the genetic covariance (covG), or heritability if trait 1 equals trait 2,

$\zeta$ is the spurious association due to bias such as population stratification, which is hypothesised to influence the test statistics consistently and irrespectively of linkage disequilibrium (in contrast to real polygenic signal).

The unweighted full LD scores for all variants are defined as

${\lambda'}=\boldsymbol{R}^{T}\boldsymbol{R}$	(Formula 8.2)

where $\boldsymbol{R}$ is an $M_{LDSR}\times M_{LDSR}$ matrix containing the standardised genotype values for each variant^40^.

The full LD scores are typically not used due to their sparseness and size and are, such as in the original LDSC software and in the Genomic SEM implementation of LDSR, instead approximated by:

${l'}_{j}=\sum_{m\in M_{window} , m\neq j} r_{j,m}^{2}$	(Formula 8.3)

which is approximating the diagonal of ${\lambda'}$.

Here  ${l'}_{j}$ is the LD score of variant $j$, obtained from the sum of genotype correlations which is the sum of the squared Pearson correlations $r_{j,m}^{2}$ between the genotypes of variant $j$ and $m$,  for the set of variants $M_{window}$ in a window defined by recombination distance, number of variants, or similar (rather than from the genome-wide set of $M_{LDSR}$), using information from a reference panel of individual level genotype or sequencing data.

In standard LDSR (recall that we refer to the implementation in Genomic SEM as the standard implementation, as this was what we base our extensions on) the LD score is further capped to a minimum of one:

${l'}_{j}=\left\{ \begin{aligned} \sum_{m\in M_{window} , m\neq j} r_{j,m}^{2}\geq1:\sum_{m\in M_{window} , m\neq j} r_{j,m}^{2} \\ \sum_{m\in M_{window} , m\neq j} r_{j,m}^{2}<1:1 \end{aligned} \right.$	(Formula 8.4)

##### 1.8.2 Weighting in standard LDSR

In standard LDSR the weighted LD scores and GWAS association Z statistics are obtained through:

$w_{aj}=\sqrt{w_{het,a,j}w_{oc,a,j}}$	(Formula 8.5)

$l_{aj}=w_{aj}{l'}_{j}$	(Formula 8.6)

$Z_{aj}=w_{aj}{Z'}_{aj}$	(Formula 8.7)

Here

$a=1,2$ for each of the two traits in the trait combination

$w_{het,a,j}$ is the heteroskedasticity correction weight

$w_{oc,a,j}$ is the correlation correction weight

${l'}_{j}$ is the unweighted LD score

${Z'}_{aj}$ is the unweighted association test Z statistic

Specifically, the correlation correction weight is obtained through:

$w_{oc,a,j}=\frac{1}{{l'}_{j}}$	(Formula 8.8)

##### 1.8.3 Overview of LDSC++

To enable more precise covG estimation from GWAS summary statistics, we extend the LDSR method based on the multivariate implementation included in Genomic SEM. We have named this extension LDSC++ (supplementary tab. S3). We validate LDSC++, first using the REML covG estimates as reference in the case of simulated traits, and second by analysing selected real world GWAS summary statistics, alongside the original LDSC method and the multivariate LDSR routine from Genomic SEM. For validations of our method extensions, standard LDSR was emulated in our LDSC++ code to additionally enforce a harmonised ordering of variants and filtering of duplicate variants across name (rs-id typically). The extensions to LDSR are:

**Extension 1 - Variable block-count:** Allowing the number of blocks to vary for when estimating covG and covG standard error, in contrast to a fixed number of blocks. Note that a variable number of blocks only influences the estimation of covG standard error when using block jackknife resampling as in original LDSR.

**Extension 2 - Extended block definitions to allow for variable block-count:** Block definitions to be used while allowing the number of blocks to vary with Extension 1. These are based on either a set number of variants or a recombination distance in cM.

**Extension 3 - Variable block-count sampling:** A novel approach to simply sample across genome blocks to estimate genome-wide covG and covG variance, rather than performing block jackknife resampling.

**Extension 4 - Extended weighting scheme for imputation quality:** An addition to the standard LDSR weighting scheme to also control for imputation quality.

- **Extension 5 - Adjusted weighting scheme to correct for the correlation between LD score and association statistic:** We adjusted the standard weighting scheme in LDSR to correct for correlation between the regression variables, to allow for LD scores smaller than one while preventing influences by extreme LD (close to zero) values due to noise.

Each extension is described in detail below.

##### 1.8.4 Extension 1 - Variable block-count

In standard LDSR, covG is estimated from one genome-wide regression while a fixed number of 200 blocks as default is used to generate covG standard errors. This is irrespective of the number of variants participating in the trait or trait pair analysis, which may vary from a few hundred thousand variants when using smaller GWASs and LD score panels to close to ten million variants for current larger GWASs and larger LD score panels. To allow the LDSR method to discriminate between situations of varying number of contributing genetic variants (Fig. 1), we made LDSC++ use a variable number of blocks rather than a fixed count. The varying number of blocks are then reflected in any associated statistical tests, where they influence the formulas for sampling variance of covG (supplementary information 5-9). Allowing the number of blocks to vary is compatible with any block definition and any sampling or resampling method used; i.e. it can be used with either number-of-variants defined blocks or recombination distance defined blocks (Extension 2), and then with either variable block-count sampling (Extension 3) or block jackknife resampling. However, for block jackknife resampling, the change only influences the estimation of standard errors.

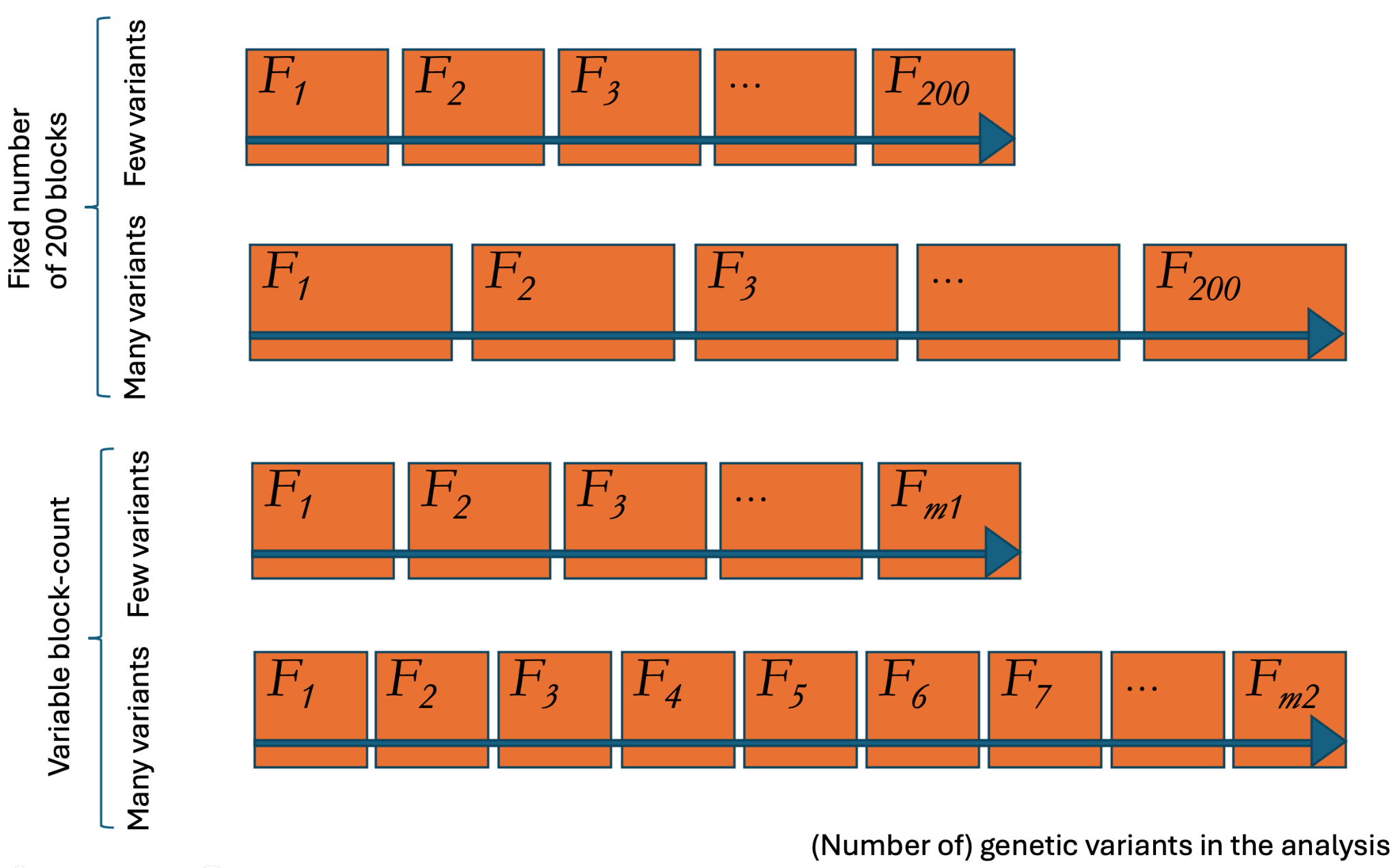

**Figure 1. Variable block-count**

A conceptual illustration of how the number of blocks for the sampling or resampling procedure vary in count proportional to the number of genetic variants in the LD score regression analysis when using LDSC++ variable block-count. The upper section represents a situation when the number of blocks are fixed to 200 and do not change in proportion to the participating number of genetic variants (as in standard LD score regression). The lower section represents a situation when allowing the number of blocks to vary (our variable block-count extension to LD score regression). For the variable block-count case, the bottom row has a larger number of variants and thus also proportionally a larger number of blocks (*m2 > m1*). To achieve this the blocks can have a defined size in terms of either number of variants/markers or recombination distance (cM). Compared to the situation with a fixed number of blocks, both *m1* and *m2* can be either larger or smaller than this arbitrary fixed number, but are displayed above as if *m2 > 200 = m1*.

##### 1.8.5 Extension 2 - Extended block definitions to allow for variable block-count

We implemented two new block definitions to allow for using a variable number of blocks (Extension 1), rather than a fixed number. These block definitions can be used with either sampling or resampling method. Briefly, our block definitions split variants into blocks based on a defined block size in terms of either 1.) number of variants or 2.) recombination distance, as opposed to blocks of varying size but a fixed variant-count as used in standard LDSR. Standard LDSR creates its blocks based on an equal number of variants, which makes it more similar to our first block definition.

For the number-of-variants defined blocks setting the variants are selected into blocks in the order of genetic coordinates and blocks will not represent the same location in the genome across traits, similar to the original LDSR block definition. For the recombination distance defined blocks setting, blocks match variants from the same positions in the genome across all traits, only including blocks that are sufficiently complete (defined as containing at least 5000 variants as default).

Both the size and count of blocks directly influences the covG standard errors obtained from the sampling or resampling method used in the LDSR. We set the default number-of-variants defined block size to 20,000 variants, for two reasons. First, this number of variants yields sufficiently small blocks for differences in number of variants across trait pairs in LDSR to be detected. Second, this number of variants, when using the 1kG3 reference or a similar reference with the same number of variants and coverage (i.e. the GLAD+ reference), conserves the independence of the per-block covG variables for the assumptions of LDSR to be valid. We aimed to keep block size as small as possible while also keeping blocks independent. The setting of 20,000 variants was validated using our selection of GWAS summary statistics (see Results).

##### 1.8.6 Extension 3 - Variable block-count sampling

To model covG heterogeneity across the genome, we implemented a new LDSR sampling method; *variable block-count sampling*. In variable block-count sampling, LDSR is performed on the genetic variant associations and LD scores contained by each of the blocks, with both the genome-wide covG and its variance being estimated from the average covG across blocks. This differs from the block jackknife resampling method, where LDSR first is performed on the full sample to obtain covG and only standard errors then are obtained by performing regressions again per block and subtracting the results from the primary regression result. We hypothesise that variable block-count sampling, as an example of segmented regression, may provide improved estimators of covG and its variance compared to a genome-wide single regression, in the presence of either non-linear relationships, heteroskedasticity, and extreme outliers. The main assumption made for the variable block-count sampling is that the within-genome heteroskedasticity of the covG variance can be ignored for the estimation of standard errors, when performing segmented regression (See the supplementary information for a detailed description of variable block-count sampling and a conceptual comparison with the block jackknife resampling method).

##### 1.8.7 Extension 4 and 5 - Extended weighting schemes for imputation quality and to correct for the correlation between LD score and association statistic

We extended the pre-existing weighting scheme in LDSR to additionally include weight components for genotype imputation quality (i.e. the INFO score) and GWAS summary statistics imputation quality (i.e. the SINFO score). The scores are here used directly as weights, as if denoting probabilities, relying on their defined range being on a scale from 0 to 1. This extension is comparable to the information score used in the LDAK^41^ heritability model. See the supplementary information for a detailed description of the extended weighting scheme in LDSC++.

A correlation correction that includes the LD score in the denominator is used in standard LDSR. To avoid division by zero, standard LDSR caps the LD score to one before weighting (see XXXX). We extended the formula for the correlation correction weights to include an extra 1 in the denominator, compared to the implementation in Genomic SEM, to allow for LD scores between zero and one. This setting is further suited to the typical range of LD scores. Our LD score panel based on the 1kG3 sample had an average LD score of about 70 for variants with MAF>0.1 (supplement S5), for which our formula will give almost indistinguishable results as compared to the original formula.

First, we allow for the LD score to fall between one and zero in addition to larger values:

${l'}_{j}=\left\{ \begin{aligned} \sum_{m\in M_{window} , m\neq j} r_{j,m}^{2}\geq0:\sum_{m\in M_{window} , m\neq j} r_{j,m}^{2} \\ \sum_{m\in M_{window} , m\neq j} r_{j,m}^{2}<0:0 \end{aligned} \right.$	(Formula 8.9)

Then, we propose the following extension to the LDSR weighting scheme as part of LDSC++:

$\dot{w}_{aj}=\sqrt{w_{het,a,j}\dot{w}_{oc,a,j}\dot{w}_{INFO,a,j}\dot{w}_{SINFO,a,j}}$	(Formula 8.10)

$l_{aj}=\dot{w}_{aj}{l'}_{j}$	(Formula 8.11)

$Z_{aj}=\dot{w}_{aj}{Z'}_{aj}$	(Formula 8.12)

Here

$a=1,2$ for each of the two traits in the trait combination

$w_{het,a,j}$ is the heteroskedasticity correction weight (unmodified)

$\dot{w}_{oc,a,j}$ is the proposed new correlation correction weight

$\dot{w}_{INFO,a,j}$ is the proposed genotype imputation uncertainty correction weight

$\dot{w}_{SINFO,a,j}$ is the proposed GWAS summary statistics imputation uncertainty correction weight

${l'}_{j}$ is the unweighted LD score

${Z'}_{aj}$ is the unweighted association test Z statistic

The proposed new correlation correction weight is obtained through:

$\dot{w}_{oc,a,j}=\frac{1}{{1+l'}_{j}}$	(Formula 8.13)

The two proposed imputation uncertainty correction weights are set to equal the imputation quality measurements for the current imputed variant, which are scores ranging from 0 to 1; the INFO score for genotype imputation and the SINFO score for GWAS summary statistics imputation, where the SINFO score is a score created to be reminiscent to the INFO score with the same range. If the variant is not imputed, these weights are set to 1.

#### 1.9 Probability distributions for LDSR

##### 1.9.1 General

We formulated simple parametric statistical tests specifically accounting for the novel varying number of blocks in the sampling and resampling methods. Second, we accounted for the fact that compared variables may not be fully independent, as is the case when comparing results from LDSR repeatedly performed on the same trait pair combination. We created tests for 1.) covG significance, 2.) covG variance significance, 3.) significance of the difference between two covG, and 4.) significance of the difference between two covG variances (See the supplementary information for a detailed description of our suggested statistical tests for LDSR).

##### 1.9.2 The estimator for block slopes as used in LDSR

LDSR is performed by dividing variant data into a number of blocks, with each variant $j$ belonging to one block $b=1\ldots B$. For standard LDSR $B=1$ for the estimation of genome-wide covG, but $B>1$ for estimation of covG standard errors (SEs; estimated via block jackknife). For LDSC++ variable block count sampling, covG and its standard errors are both calculated using the same value of $B$, with $B>1$ .

In standard LDSR, for covG SE estimation, the number of blocks is set to 200 as default, and a block is defined accordingly as to have roughly equal number of variance in each block, and variants ordered as they appear in the reference file. In LDSC++ we instead define blocks based on our novel number-of-variants or recombination distance block-definitions.

Using Form. 8.1, the slope parameter per block is:

$\beta_{gb}=\frac{\sqrt{N_{1}N_{2}}}{M_{LDSR}}\rho_{gb}$	(Formula 9.1)

In LDSR, $\beta_{gb}$ is estimated by maximum likelihood (ML) or ordinary least squares (OLS)^42^ when regressing $Z_{1j}Z_{2j}$ on $l_{j}$. Therefore, the estimator of the per-block slope has the distribution:

$\hat{\beta}_{gb}\sim N\left( \beta_{gb},\sigma_{\hat{\beta}_{gb}}^{2} \right)$	(Formula 9.2)

where

$\sigma_{\hat{\beta}_{gb}}^{2}=\frac{\sigma_{b}^{2}}{\sum_{j \in b} \left( l_{j}-\bar{l} \right)^{2}}$	(Formula 9.3)

Here $\sigma_{b}^{2}$ is the variance of the residuals in the simple linear regression model.

Using the distribution of $\hat{\beta}_{gb}$ (Form 9.2) and Form. 9.1, it can be shown that the estimator $\hat{\rho}_{gb}$ of the block-wise genetic covariance $\rho_{gb}$ has the distribution:

$\hat{\rho}_{gb}\sim N\left( \rho_{gb},\sigma_{\hat{\rho}_{gb}}^{2} \right)$	(Formula 9.4)

where

$\rho_{gb}=\frac{M_{LDSR}}{\sqrt{N_{1}N_{2}}}\beta_{gb}$	(Formula 9.5)

and

$\sigma_{\hat{\rho}_{gb}}^{2}=\frac{{{(M}_{LDSR})}^{2}}{N_{1}N_{2}}\sigma_{\hat{\beta}_{gb}}^{2}=\frac{{{(M}_{LDSR})}^{2}}{N_{1}N_{2}}\frac{\sigma_{b}^{2}}{\sum_{j \in b} \left( l_{j}-\bar{l} \right)^{2}}$	(Formula 9.6)

Depending on different assumptions made, such as in the two different types of estimators for genome-wide covG and its variance that we describe further on, a block can be defined to cover the whole genome or only a subset.

1.9.3 A maximum likelihood estimator for variance of block slopes in LDSR

Looking within block, using a conditional normal regression model approximation of a bivariate normal situation, and hence treating the LD scores as constants and only the association statistics as random, an unbiassed maximum likelihood (ML) estimator of $\sigma_{\hat{\beta}_{gb}}^{2}$ is^42^:

$S_{\hat{\beta}_{gb}}^{2}=\frac{M_{b}}{M_{b}-2}\sigma_{b}^{2}$	(Formula 9.7)

which follows distribution^42^:

$S_{\hat{\beta}_{gb}}^{2}\sim\frac{\sigma_{b}^{2}}{\left( M_{b}-2 \right)} \chi^{2}\left( M_{b}-2 \right)$	(Formula 9.8)

Here, $M_{b}$ is the number of variants in the current block. The estimator $S_{\hat{\beta}_{gb}}^{2}$ approaches a normal distribution as $M_{b}\to\infty$. Since $M_{b}$ usually is very large, usually >> 200 variants, we may assume normality in our case. See 1.9.6 Normal approximation of covG variance estimators below.

In the same way as for the ML/OLS covG estimator, depending on different assumptions made, such as in the two different types of estimators for genome-wide covG and its variance that we describe further on, a block can be defined to cover the whole genome or only a subset.

##### 1.9.4 An estimator for standard LDSR

Standard LDSR relies on the ML/OLS estimator for estimating the slope and thus covG (Form. 9.2, 9.4), and then applies block jackknife resampling to estimate the variance of the same.

Standard LDSR makes the assumptions that:

1.) There is one $\beta_{g}$ and $\rho_{g}$ with genome-wide variances to be estimated and,
2.) Block variables are independent across blocks

In standard LDSR, the slope is estimated from one single regression for the whole genome as:

$\hat{\beta}_{g,B=1}= \hat{\beta}_{gb}\left[ b=1=B \right]$	(Formula 9.9)

The variance of this genome-wide slope is not estimated from the regression residuals (Form. 9.3 and 9.6) directly, but rather through block jackknife resampling using a default of 200 blocks. Further, the slope from the genome-wide one-block regression $\hat{\beta}_{g,B=1}$is used both as the estimate of the genome-wide slope (rather than the average over jackknife pseudovalues $\hat{\beta}_{g,jack}$ ), as the estimate of the full data when computing pseudovalues and in the formula for the variance of the genome-wide slope estimator.

That is, in the block jackknife^43,44^, we calculate pseudovalues:

$\hat{\beta}_{gb,jack}=B\hat{\beta}_{g,B=1}-\left( B-1 \right)\hat{\beta}_{gb}$	(Formula 9.10)

and then use these pseudovalues to calculate:

$\hat{\beta}_{g,jack}=\frac{1}{B}\sum_{b=1}^{B} \hat{\beta}_{gb,jack}=\frac{1}{B}\sum_{b=1}^{B} (B\hat{\beta}_{g,B=1}-\left( B-1 \right)\hat{\beta}_{gb})$	(Formula 9.11)

This $\hat{\beta}_{g,jack}$ can be used to estimate the variance of $\beta_{g}$. However, for standard LDSR the block jackknife estimator of this variance is computed instead using the genome-wide slope $\hat{\beta}_{g,B=1}$ and the jackknife block estimates (pseudovalues) as:

$S_{\hat{\beta}_{g,jack}}^{2}=\frac{1}{B\left( B-1 \right)}\sum_{b=1}^{B} \left( \hat{\beta}_{gb,jack}-\hat{\beta}_{g,B=1} \right)^{2}$	(Formula 9.12)

Then:

$\hat{\rho}_{g,B=1}=\frac{M_{LDSR}}{\sqrt{N_{1}N_{2}}}\hat{\beta}_{g,B=1}$	(Formula 9.13)

$\hat{\rho}_{g,jack}=\frac{M_{LDSR}}{\sqrt{N_{1}N_{2}}}\hat{\beta}_{g,jack}=\frac{M_{LDSR}}{\sqrt{N_{1}N_{2}}}\frac{1}{B}\sum_{b=1}^{B} \hat{\beta}_{gb,jack}$	(Formula 9.14)

$S_{\hat{\rho}_{g,jack}}^{2}=\frac{{{(M}_{LDSR})}^{2}}{N_{1}N_{2}}S_{\hat{\beta}_{g,jack}}^{2}=\frac{{{(M}_{LDSR})}^{2}}{N_{1}N_{2}}\frac{1}{B\left( B-1 \right)}\sum_{b=1}^{B} \left( \hat{\beta}_{gb,jack}-\hat{\beta}_{g,B=1} \right)^{2}$	(Formula 9.15)

The distribution of $\hat{\beta}_{g,B=1}$ and $\hat{\rho}_{g,B=1}$ can be obtained from the block-wise distributions above. We have not found reliable information on the exact distributions of block jackknife estimators, but assume that each $\hat{\beta}_{gb,jack}$ is part of a random sample- that is each $\hat{\beta}_{gb,jack}$ is independent and identically distributed (IID), such that:

$\hat{\beta}_{g,jack}\sim N\left( \beta_{g}, \sigma_{\hat{\beta}_{g,jack}}^{2} \right)$	(Formula 9.16)

Assuming that there is such as population parameter as $\beta_{g}$ with variance $\sigma_{\beta_{g}}^{2}$, the estimator $S_{\hat{\beta}_{g,jack}}^{2}$ can be thought of having the distribution of an unbiassed estimator of a sample variance:

$S_{\hat{\beta}_{g,jack}}^{2}\sim\frac{\sigma_{\hat{\beta}_{g,jack}}^{2}}{B-1}\chi^{2}\left( B-1 \right)$	(Formula 9.17)

Note again that $\hat{\beta}_{g,jack}$ is not used as output from the standard LDSR, which instead yields $\hat{\beta}_{g,B=1}$ as part of its final estimate towards the genome-wide covG.

Intercepts are estimated analogous to how the slopes are estimated. For standard LDSR the intercept is obtained from the genome-wide one-block regression intercept.

##### 1.9.5 An estimator for LDSC++ variable block-count sampling

We created LDSC++ to extend the existing LDSR method, not be revolutionary different from it, and to be able to produce recognisable results. While our variable block-count sampling aims to acknowledge heterogeneity in local heritability and LD patterns across the genome, it still assumes, like standard LDSR, that there are meaningful constructs represented by genome-wide population parameters.

We do however expect an improvement from using our sampling approach as our estimators make use of the sample average and effectively implements a segmented regression (with much potential for improvement) rather than one genome-wide regression.

The statistical tests we have set up here for variable block-count sampling are relying on the same assumptions made for standard LDSR about:

1.) There is one $\beta_{g}$ and $\rho_{g}$ with genome-wide variances to be estimated and,
2.) Block variables are independent across blocks.

The difference between variable block-count sampling and default LDSR block jackknife resampling, apart from allowing the number of blocks to vary, is that we consider the slope, and so genetic covariance, for each block to form part of a random sample across the blocks. We then estimate a genome-wide slope and variance using the sample mean and sample variance based on an equal number of blocks > 1:

$\hat{\beta}_{g,vbcs}=\frac{1}{B}\sum_{b=1}^{B} \hat{\beta}_{gb}$	(Formula 9.18)

$S_{\hat{\beta}_{g,vbcs}}^{2}=\frac{1}{B-1}\sum_{b=1}^{B} \left( \hat{\beta}_{gb}-\hat{\beta}_{g,vbcs} \right)^{2}$	(Formula 9.19)

Then:

$\hat{\rho}_{g,vbcs}=\frac{M_{LDSR}}{\sqrt{N_{1}N_{2}}}\hat{\beta}_{g,vbcs}=\frac{M_{LDSR}}{\sqrt{N_{1}N_{2}}}\frac{1}{B}\sum_{b=1}^{B} \hat{\beta}_{gb}=\frac{1}{B}\sum_{b=1}^{B} \hat{\rho}_{gb}$	(Formula 9.20)

$S_{\hat{\rho}_{g,vbcs}}^{2}=\frac{{{(M}_{LDSR})}^{2}}{N_{1}N_{2}}S_{\hat{\beta}_{g,vbcs}}^{2}=\frac{{{(M}_{LDSR})}^{2}}{N_{1}N_{2}}\frac{1}{B-1}\sum_{b=1}^{B} \left( \hat{\beta}_{gb}-\hat{\beta}_{g,vbcs} \right)^{2}$	(Formula 9.21)

As the LDSC++ variable block-count sampling is a simple random sample, where the individual block estimates can be assumed to have known distribution properties, the random distributions of the genome-wide estimators are well known. The average covG across blocks equalling the variable block-count sampling estimator, is:

$\hat{\beta}_{g,vbcs}=\frac{1}{B}\sum_{b=1}^{B} \hat{\beta}_{gb}\sim N\left( \beta_{g},\sigma_{\hat{\beta}_{g,vbcs}}^{2} \right)$	(Formula 9.22)

Where $S_{\hat{\beta}_{g,vbcs}}^{2}$ is the unbiassed estimator for the sample variance, with distribution:

$$S_{\hat{\beta}_{g,vbcs}}^{2} \sim\frac{\sigma_{\hat{\beta}_{g,vbcs}}^{2}}{(B-1)} \chi^{2}(B-1)$$

(Formula 9.23)

##### 1.9.6 Normal approximation of covG variance estimators

The covG sampling variance estimators from either method can be approximated by a normal as the degrees of freedom gets large:

$S_{\hat{\beta}_{g,any}}^{2}\to\frac{\sigma_{\hat{\beta}_{g,any}}^{2}}{(B-1)}N\left( B-1, 2\left( B-1 \right) \right)$	(Formula 9.24)

$$\Longleftrightarrow$$

$S_{\hat{\beta}_{g,any}}^{2}\to\sigma_{\hat{\beta}_{g,any}}^{2}N\left( 1, \frac{2}{B-1} \right)$	(Formula 9.25)

or

$S_{\hat{\beta}_{g,any}}^{2}\to N\left( \sigma_{\hat{\beta}_{g,any}}^{2}, \frac{2\sigma_{\hat{\beta}_{g,any}}^{4}}{B-1} \right)$	(Formula 9.26)

where $S_{\hat{\beta}_{g,any}}^{2}$ is either $S_{\hat{\beta}_{g,jack}}^{2}$ or $S_{\hat{\beta}_{g,vbcs}}^{2}$, which may have slightly different behaviours as explained below in 1.9.7 .

LDSR intercepts can be estimated analogous to how the slopes are estimated. For LDSC++ variable block-count sampling the genome-wide intercept is obtained as a mean of per-block intercepts.

##### 1.9.7 The difference between the estimators for standard LDSR and LDSC++ variable block-count sampling

It is possible to show the exact difference between the two methods, LDSR default block jackknife resampling and LDSC++ variable block-count sampling, and that it is based on the difference between how the genome-wide estimates are obtained.

For the slopes:

$\hat{\beta}_{g,B=1}=\hat{\beta}_{g,vbcs}$	(Formula 9.27)

$\hat{\beta}_{g,B=1}=\frac{1}{B}\sum_{b=1}^{B} \hat{\beta}_{gb}$	(Formula 9.28)

For the variances:

$S_{\hat{\beta}_{g,jack}}^{2}=S_{\hat{\beta}_{g,vbcs}}^{2}$	(Formula 9.29)

$\frac{1}{B\left( B-1 \right)}\sum_{b=1}^{B} \left( \hat{\beta}_{gb,jack}-\hat{\beta}_{g,B=1} \right)^{2}=\frac{1}{B-1}\sum_{b=1}^{B} \left( \hat{\beta}_{gb}-\hat{\beta}_{g,vbcs} \right)^{2}$	(Formula 9.30)

$\frac{1}{B}\left( \hat{\beta}_{gb,jack}-\hat{\beta}_{g,B=1} \right)^{2}=\left( \hat{\beta}_{gb}-\hat{\beta}_{g,vbcs} \right)^{2}$	(Formula 9.31)

$\frac{\left( B-1 \right)}{B}\left( \hat{\beta}_{gb}-\hat{\beta}_{g,B=1} \right)^{2}=\left( \hat{\beta}_{gb}-\hat{\beta}_{g,vbcs} \right)^{2}$	(Formula 9.32)

For $B\to\infty$

$\hat{\beta}_{g,B=1}=\hat{\beta}_{g,vbcs}$	(Formula 9.33)

$\hat{\beta}_{g,B=1}=\frac{1}{B}\sum_{b=1}^{B} \hat{\beta}_{gb}$	(Formula 9.34)

which is the same equality as for the slopes.

This equality is true when $E\left[ \hat{\beta}_{g,B=1} \right]=E\left[ \hat{\beta}_{gb} \right]$, for all blocks. According to our earlier hypotheses, we think that this is not always the case. This is despite the distribution assumptions made and the corresponding statistical tests being reliant on the assumption that each block estimator has the same expected value as the genome-wide estimators and population parameters.

There are reasons concerning modelling for generally treating the genome-wide population parameters as averages over heterogenous genetically local parameters, as this may better reflect reality. We can think of situations when the underlying model is better represented by a segmented (piece-wise) regression as in our variable block-count sampling, rather than one genome-wide regression as in default LDSR block jackknife resampling:

In the presence of

1.) non-linear relationships
2.) heteroskedasticity
3.) extreme outliers in combination with measurement error

#### 1.10 Statistical tests for LDSR

##### 1.10.1 Test of covG

The default test of LDSR covG significance has previously been made using the LDSR estimator of covG; $\hat{\rho}_{g,B=1}$, and the corresponding block jackknife resampling estimator of covG variance; $S_{\hat{\rho}_{g,jack}}^{2}$.

We have additionally presented new estimators based on our proposed variable block-count sampling method, where we estimate covG with $\hat{\rho}_{g,vbcs}$ and covG variance with $\hat{\sigma}_{\rho_{g,vbcs}}^{2}$.

Estimators of either method (here symbolised by $any$) can be used to construct test statistics (denoted $T$ ) to test the if covG is statistically different from 0.

For $\rho_{g}=\frac{M_{LDSR}}{\sqrt{N_{1}N_{2}}}\beta_{g}$:

$H_{0}: \beta_{g}=0$

$H_{1}: \beta_{g}\neq0$

$T_{\hat{\rho}_{g,any}}=\hat{\rho}_{g,any}= \frac{M_{LDSR}}{\sqrt{N_{1}N_{2}}}\hat{\beta}_{g,any}$	(Formula 10.1)

following the distribution:

$T_{\hat{\rho}_{g,any}}\sim\frac{M_{LDSR}}{\sqrt{N_{1}N_{2}}}N\left( 0,\frac{\sigma_{\hat{\beta}_{g,any}}^{2}}{B} \right)$	(Formula 10.2)

Using $BS_{\hat{\beta}_{g,any}}^{2}$ to infer $\sigma_{\hat{\beta}_{g,any}}^{2}$ , and a two-sided test.

Then we obtain the test of covG significance implemented in default Genomic SEM LDSR.

##### 1.10.2 Test of covG variance

To the best of our knowledge there has not been any default test of LDSR covG variance significance in any previous implementation of LDSR. It is however possible to perform such a test using the default LDSR block jackknife estimator $S_{\hat{\rho}_{g,jack}}^{2}$.We have additionally presented a new estimator based on our proposed variable block-count sampling method, where we estimate covG variance with $S_{\hat{\rho}_{g,vbcs}}^{2}$.

Estimators of either method (here symbolised by $any$), can be used to construct test variables to test the significance of covG variance.

For $\sigma_{\hat{\rho}_{g,any}}^{2}=\frac{{{(M}_{LDSR})}^{2}}{N_{1}N_{2}}\sigma_{\hat{\beta}_{g,any}}^{2}$:

$H_{0}: \frac{{{(M}_{LDSR})}^{2}}{N_{1}N_{2}}\sigma_{\hat{\beta}_{g,any}}^{2}=c$

$H_{1}: \frac{{{(M}_{LDSR})}^{2}}{N_{1}N_{2}}\sigma_{\hat{\beta}_{g,any}}^{2}>c$

$T_{S_{\hat{\rho}_{g,any}}^{2}}=\frac{{{(M}_{LDSR})}^{2}}{N_{1}N_{2}}S_{\hat{\beta}_{g,any}}^{2}\sim\frac{{{(M}_{LDSR})}^{2}}{N_{1}N_{2}}{\frac{\sigma_{\hat{\beta}_{g,any}}^{2}}{(B-1)} \chi}^{2}(B-1)$	(Formula 10.3)

Using $BS_{\hat{\beta}_{g,any}}^{2}$ to infer $\sigma_{\hat{\beta}_{g,any}}^{2}$, $c$ being an arbitrary constant $c\neq0$ , and a one-sided test.

It should be possible to obtain useful values for the constant $c$ based on empirical data.

##### 1.10.3 Test of difference between two covG

The test of the difference between two covG is set up to test whether the difference in two estimates of covG would indicate either 1.) a difference in population parameters (i.e. hypothesis 1 below) when comparing estimates made on different putative covG, or 2.) that the recorded difference is due to a difference in estimator performance when comparing results on the same covG but using different estimators, as we used to compare the performance of our estimators.

For the sake of testing the differences between results of our different methods and estimators, we compare the results from each method directly, motivated by our project data on estimated covG and their standard errors which seem to follow extremely similar distributions irrespective of method.

For the test we have two genetic covariance population parameters $\rho_{g}\left( 1 \right)$ and $\rho_{g}\left( 2 \right)$ with two estimates $\hat{\rho}_{g,any}\left( 1 \right)$ and $\hat{\rho}_{g,any}\left( 2 \right)$, using number of blocks $B(1)$ and $B(2)$, coming from either the LDSR method or our updated estimator.

For $\rho_{g}(1)=\frac{M_{LDSR}}{\sqrt{N_{1}N_{2}}}\beta_{g}(1)$ and $\rho_{g}(2)=\frac{M_{LDSR}}{\sqrt{N_{1}N_{2}}}\beta_{g}(2)$:

$H_{0}: \beta_{g}\left( 1 \right)=\beta_{g}\left( 2 \right)$

$H_{1}: \beta_{g}\left( 1 \right)\neq\beta_{g}\left( 2 \right)$

We first construct a sample test-variable of the mean block covG for each of the two estimates; $R_{\hat{\rho}_{g,any}\left( 1 \right)}$ and $R_{\hat{\rho}_{g,any}\left( 2 \right)}$ , with corresponding test variables for the same difference on the scale of LDSR slopes; $R_{\hat{\beta}_{g,any}\left( 1 \right)}$ and $R_{\hat{\beta}_{g,any}\left( 2 \right)}$. The test variables have the form and distribution as in Form. 10.1 and 10.2 from the previous test of covG significance. The notation is to make it clear that they represent test statistics with the accompanying distribution, as opposed to estimators of population parameters.

Then the test variable for the difference of the samples is:

$T_{\Delta\hat{\rho}_{g,any}}=R_{\hat{\rho}_{g,any}\left( 1 \right)}- R_{\hat{\rho}_{g,any}\left( 2 \right)}=\frac{M_{LDSR}}{\sqrt{N_{1}N_{2}}}\left( R_{\hat{\beta}_{g,any}(1)}- R_{\hat{\beta}_{g,any}(2)} \right)$	(Formula 10.4)

$T_{\Delta\hat{\rho}_{g,any}}\sim\frac{M_{LDSR}}{\sqrt{N_{1}N_{2}}} N\left( \beta_{g}\left( 1 \right)-\beta_{g}(2),\sigma_{R_{\hat{\beta}_{g,any}(1)}- R_{\hat{\beta}_{g,any}(2)}}^{2} \right)$	(Formula 10.5)

The variance of the difference between the test variables on the scale of the slope is:

$\sigma_{R_{\hat{\beta}_{g,any}(1)}- R_{\hat{\beta}_{g,any}(2)}}^{2}=\frac{\sigma_{\hat{\beta}_{g,any}\left( 1 \right)}^{2}}{B(1)}+\frac{\sigma_{\hat{\beta}_{g,any}\left( 2 \right)}^{2}}{B(2)}-2Cov\left( R_{\hat{\beta}_{g,any}(1)},R_{\hat{\beta}_{g,any}(2)} \right)$	(Formula10.6)

$=\frac{\sigma_{\hat{\beta}_{g,any}\left( 1 \right)}^{2}}{B(1)}+\frac{\sigma_{\hat{\beta}_{g,any}\left( 2 \right)}^{2}}{B(2)}-2\frac{\sigma_{\hat{\beta}_{g,any}\left( 1 \right)}}{\sqrt{B(1)}}\frac{\sigma_{\hat{\beta}_{g,any}\left( 2 \right)}}{\sqrt{B(2)}}Corr\left( R_{\hat{\beta}_{g,any}(1)},R_{\hat{\beta}_{g,any}(2)} \right)$	(Formula 10.7)

For this test we use $B(1)S_{\hat{\beta}_{g,any}}^{2}(1)$ to infer $\sigma_{\hat{\beta}_{g,any}}^{2}(1)$ and the equivalent procedure for the second parameter, and a two-sided test as the distribution is symmetrical.

The covariance between the variables can be estimated from the data if measurements of LDSR $\hat{\beta}_{gb}$ are available, or if the covariance is otherwise known.

For highly correlated variables, such as the results of two slightly different methods applied on the same trait combination, we propose that when $Corr\left( R_{\hat{\beta}_{g,any}\left( 1 \right)},R_{\hat{\beta}_{g,any}\left( 2 \right)} \right)\to1$ then the variance of the difference between the test variables on the scale of the slope can be approximated as:

$$\sigma_{R_{\hat{\beta}_{g,any}\left( 1 \right)}- R_{\hat{\beta}_{g,any}\left( 1 \right)}}^{2}\approx\frac{\sigma_{\hat{\beta}_{g,any}\left( 1 \right)}^{2}}{B(1)}+\frac{\sigma_{\hat{\beta}_{g,any}\left( 2 \right)}^{2}}{B(2)}-2\frac{\sigma_{\hat{\beta}_{g,any}\left( 1 \right)}}{\sqrt{B(1)}}\frac{\sigma_{\hat{\beta}_{g,any}\left( 2 \right)}}{\sqrt{B(2)}}$$

$=\left( \frac{\sigma_{\hat{\beta}_{g,any}\left( 1 \right)}}{\sqrt{B(1)}}-\frac{\sigma_{\hat{\beta}_{g,any}\left( 2 \right)}}{\sqrt{B(2)}} \right)^{2}$	(Formula 10.8)

##### 1.10.4 Test of difference between the variances of two covG

The test of the difference between two covG variances is set up to test whether the difference in two estimates of covG variance would indicate either 1.) a difference in population parameters (i.e. hypothesis 1 below) when comparing estimates made on different putative covG, or 2.) that the recorded difference is due to a difference in estimator performance when comparing results on the same covG but using different estimators, as we used to compare the performance of our estimators.

For the test we have two genetic covariance population parameters $\rho_{g}\left( 1 \right)$ and $\rho_{g}\left( 2 \right)$ with two estimates $\hat{\rho}_{g,any}\left( 1 \right)$ and $\hat{\rho}_{g,any}\left( 2 \right)$, using number of blocks $B(1)$ and $B(2)$, coming from either LDSR method or estimator. These two estimates have a distribution, with variances $\sigma_{\hat{\rho}_{g,any}(1)}^{2}$ and $\sigma_{\hat{\rho}_{g,any}(2)}^{2}$ respectively.

For $\sigma_{\hat{\rho}_{g,any}(1)}^{2}=\frac{{{(M}_{LDSR})}^{2}}{N_{1}N_{2}}\sigma_{\hat{\beta}_{g,any}(1)}^{2}$ and $\sigma_{\hat{\rho}_{g,any}(2)}^{2}=\frac{{{(M}_{LDSR})}^{2}}{N_{1}N_{2}}\sigma_{\hat{\beta}_{g,any}(2)}^{2}$:

$H_{0}: \sigma_{\hat{\beta}_{g,any}(1)}^{2}=\sigma_{\hat{\beta}_{g,any}(2)}^{2}=c$

$H_{1}: \sigma_{\hat{\beta}_{g,any}(1)}^{2}\neq\sigma_{\hat{\beta}_{g,any}(2)}^{2}$

Recall that an approximate distribution, using the normal approximation, for $S_{\hat{\rho}_{g,any}}^{2}$ is (Form. 9.24 – 9.26):

$S_{\hat{\rho}_{g,any}}^{2}=\frac{{{(M}_{LDSR})}^{2}}{N_{1}N_{2}}S_{\hat{\beta}_{g,any}}^{2} \sim\frac{{{(M}_{LDSR})}^{2}}{N_{1}N_{2}}N\left( \sigma_{\hat{\beta}_{g,any}}^{2}, \frac{2\sigma_{\hat{\beta}_{g,any}}^{4}}{B-1} \right)$

The test variable describing the approximate distribution for the difference in estimated variances can therefore be set up as:

$T_{\Delta S_{\hat{\rho}_{g,any}}^{2}}=S_{\hat{\rho}_{g,any}(1)}^{2}-S_{\hat{\rho}_{g,any}\left( 2 \right)}^{2}=\frac{{{(M}_{LDSR})}^{2}}{N_{1}N_{2}}\left( S_{\hat{\beta}_{g,any}(1)}^{2}-S_{\hat{\beta}_{g,any}(2)}^{2} \right)$	(Formula 10.9)

$T_{\Delta S_{\hat{\rho}_{g,any}}^{2}}\sim\frac{{{(M}_{LDSR})}^{2}}{N_{1}N_{2}}N\left( \begin{matrix} \sigma_{\hat{\rho}_{g,any}(1)}^{2}-\sigma_{\hat{\rho}_{g,any}(2)}^{2}, & Var\left( S_{\hat{\beta}_{g,any}(1)}^{2}-S_{\hat{\beta}_{g,any}(2)}^{2} \right) \end{matrix} \right)$	(Formula 10.10)

where:

$Var\left( S_{\hat{\beta}_{g,any}(1)}^{2}-S_{\hat{\beta}_{g,any}(2)}^{2} \right)=\frac{2\sigma_{\hat{\beta}_{g,any}(1)}^{4}}{B(1)-1}+\frac{2\sigma_{\hat{\beta}_{g,any}(2)}^{4}}{B(2)-1}-2Cov\left( S_{\hat{\beta}_{g,any}(1)}^{2},S_{\hat{\beta}_{g,any}(2)}^{2} \right)=\frac{2\sigma_{\hat{\beta}_{g,any}(1)}^{4}}{B\left( 1 \right)-1}+\frac{2\sigma_{\hat{\beta}_{g,any}(2)}^{4}}{B\left( 2 \right)-1}-2\sqrt{\frac{2\sigma_{\hat{\beta}_{g,any}(1)}^{4}}{B\left( 1 \right)-1}}\sqrt{\frac{2\sigma_{\hat{\beta}_{g,any}(2)}^{4}}{B\left( 2 \right)-1}}Corr\left( S_{\hat{\beta}_{g,any}(1)}^{2},S_{\hat{\beta}_{g,any}(2)}^{2} \right)$	(Formula 10.11)

The test is then performed using $B(1)S_{\hat{\beta}_{g,any}}^{2}(1)$ to infer $\sigma_{\hat{\beta}_{g,any}}^{2}(1)$ and the equivalent procedure for the second parameter, and a two-sided test.

In the same way as for covG differences, the covariance between the variables can be estimated from the data if measurements of LDSR $\hat{\beta}_{gb}$ are available, or if the covariance is otherwise known.

Also, similar to the approximation for covG differences, for highly correlated variables, such as the results of two slightly different methods applied on the same trait combination, we propose that when $Corr\left( S_{\hat{\beta}_{g,any}(1)}^{2},S_{\hat{\beta}_{g,any}(2)}^{2} \right)\to1$ then the variance of the difference between the test variables on the scale of the slope can be approximated as:

$Var\left( S_{\hat{\beta}_{g,any}(1)}^{2}-S_{\hat{\beta}_{g,any}(2)}^{2} \right)\approx\frac{2\sigma_{\hat{\beta}_{g,any}(1)}^{4}}{B\left( 1 \right)-1}+\frac{2\sigma_{\hat{\beta}_{g,any}(2)}^{4}}{B\left( 2 \right)-1}-2\sqrt{\frac{2\sigma_{\hat{\beta}_{g,any}(1)}^{4}}{B\left( 1 \right)-1}}\sqrt{\frac{2\sigma_{\hat{\beta}_{g,any}(2)}^{4}}{B\left( 2 \right)-1}}=\left( \sqrt{\frac{2\sigma_{\hat{\beta}_{g,any}(1)}^{4}}{B\left( 1 \right)-1}}-\sqrt{\frac{2\sigma_{\hat{\beta}_{g,any}(2)}^{4}}{B\left( 2 \right)-1}} \right)^{2}$	(Formula 10.12)

##### 1.10.5 Complementary non-parametric test

As a complement to our proposed parametric statistical tests, we performed an additional paired samples Wilcoxon signed-rank test^45^ on the differences in covG and covG variance. The Wilcoxon signed-rank test is a non-parametric test that does not rely on assumptions on the underlying statistical distributions, which our other tests do. The Wilcoxon signed-rank test is also an example of a paired differences test, which assumes a high degree of correlation within the pair variables, as we expected to be the case in our comparisons. The downside is less power to detect statistically significant differences, compared to parametric alternatives (like our other proposed tests).

##### 1.10.6 Correcting for multiple comparisons

After multiple statistical tests on comparisons between covG or standard errors of covG, in matrix form, p-values were adjusted for multiple comparisons using the Benjamini & Hochberg^46^ method (q-values), which is also called false discovery rate (FDR). The effective number of comparisons for the correction was set to the number of principal components of the resampling covariance (i.e. the ‘V’ matrix in Genomic SEM) of the covG that would explain >99.5% of the eigenvalue variance, to obtain an estimate of the number of comparisons adjusted for the correlations between variables^47–49^. This value for effective number of comparisons was used for multiple comparison adjustments both for comparisons between covG and between their resampling standard errors. Using an effective number of comparisons in FDR that is smaller than the length of the vector of p-values requires forcing the number of tests to a specified value. This approach combines the utility of using an effective number of tests, warranted by correlation patterns in the data, with the less conservative FDR method.

### 2 Supplementary results

#### 2.1 Differences between LD score regression implementations

To detect differences associated with implementation details not related to our novel extensions, such as differences in software libraries or packages used in each program, we compared the results from three implementations of LDSR; LDSC, Genomic SEM LDSC, and LDSC++, without activating our novel extensions. Specifically, we ran these using the European 1kG3 LD scores otherwise used in the study, a MAF > 0.01 filter, an INFO > 0.6 filter, a consistent setting of 200 blocks and block jackknife resampling.

##### 2.1.1 LDSC++ compared to Genomic SEM LDSC

**covG:** In this setting LDSC++ produced on average 4% larger covG and 3.4% larger h^2^ compared to Genomic SEM LDSC. Some differences, particularly when including SCHI, were statistically significant. The Wilcoxon signed-rank test indicated significant (p=0.002) difference in covG between the methods.

**covG variance:** LDSC++ produced on average 2.7% smaller covG CV, or 4.7% smaller considering h^2^ CV only. Many of the differences were statistically significant, mostly covering the same trait combinations that showed significant differences in covG. However, the Wilcoxon signed-rank test did not indicate significant differences in covG variance between the methods.

##### 2.2.2 LDSC++ compared to LDSC

For comparisons with LDSC, we only investigated results of h^2^ (diagonals in the genomic covariance matrix) for practical reasons. The effective number of comparisons for the statistical tests was still set according to a full matrix however.

**covG:** LDSC++ produced on average 2.9% larger h^2^ compared to LDSC. The differences were significant for ANXI, BMI, MDDB, and SCHI. However, the Wilcoxon signed-rank test did not indicate significant difference in covG between the methods.

**covG variance:** LDSC++ yielded on average 2.8% smaller h^2^ CV. The differences were significant for ANXI, BIPO, BMI, MDDB, EDUA, and INSO. However, the Wilcoxon signed-rank test did not indicate significant differences in covG variance between the methods.

#### 2.2 Validation of default block size settings

As a prerequisite for using the default number-of-variants block setting of 20,000 variants per block, it was validated to avoid excessively small blocks in terms of recombination distance. We compared covG derived with variable block-count sampling and the default number-of-variants defined block setting with that of using blocks additionally constrained to a minimum recombination distance span of 5 cM. Negligible differences in results and block-count were observed across the selected datasets between these two settings, indicating that for our selection of traits and reference panel, blocks of 20,000 variants spanning less than 5 cM did not have a noticeable impact on the results.

#### 2.3 Differences between LD score regression implementations

To detect differences associated with implementation details not related to our novel extensions, such as differences in software libraries or packages used in each program, we compared the results from LDSC++ with the other two implementations of LDSR; LDSC, and Genomic SEM LDSC, without activating our novel extensions (supplementary tab. S9A). Specifically, we ran these using the European 1kG3 LD scores otherwise used in the study, a MAF > 0.01 filter, an INFO > 0.6 filter, a consistent setting of 200 blocks and block jackknife resampling. The LDSC++ implementation yielded larger covG and smaller covG estimates compared to both other methods, that can be attributable to implementation differences between programs. For example, differentially treating duplicate variants may explain some of the differences, which we would have to account for when further comparing methods.

#### 2.4 Extensions to LD score regression applied to simulated GWAS

See the main manuscript for validation results on individual level data from our GLAD+ sample.

The REML h^2^ for the simulated traits SIM1 and SIM2 were estimated in the range from 13% to 15%, and the REML r_g_ between traits in a pair to 0.43, 0.24, 0.19, and 0.20 for the QTL sharing fractions 100%, 75%, 50%, and 25% respectively (supplementary tab. S8A, S8B). Note that imputation quality was not modelled in our simulated trait GWASs for practical reasons, and the evaluation of the imputation quality weighting extension was not included in the validation performed on simulated traits. Using the REML estimates as reference, we calculated average RMSD over both traits in the trait pair and all QTL sharing fractions for covG estimates, their standard errors and their CV (supplementary tab. S8A, S8B). The default block-size settings for the number-of-variants block definition yielded a block-count of 391; equal for all bivariate trait pairs. The default recombination distance block definition yielded block-counts in the range from 338 to 363.

**covG:** The average covG estimates across all QTL sharing fractions were seen to be extremely similar between methods, even though some variability could be observed in the results for different simulated trait pairs (Fig. 2, supplementary tab. S8A, S8B). We recorded a downward average relative bias for all LDSR methods in the range from -1% to -8% compared to REML. The block jackknife resampling method (RMSE=∈[0.0103, 0.0104] | rel.Δ∈[-1%,-4%]) yielded more accurate overall average covG estimates as compared to variable block-count sampling (RMSE∈[0.0137,0.0191] | rel.Δ∈[-5%,-8%]) across block definitions and measured across all simulated traits and QTL sharing fractions, with the REML results as reference. Using the recombination distance block definition yielded consistently more accurate and less biassed covG estimates (RMSE∈[0.0103,0.0190], rel.Δ∈[-1%,-5%]) compared to when using the number-of-variants block definition (RMSE∈[0.0103,0.0138], rel.Δ∈[-3%,-8%]); especially for off-diagonal elements and when using block jackknife resampling, across (re)sampling methods and measured across all simulated traits and QTL sharing fractions.

**covG variance:** We recorded a predominantly downward relative bias in covG CV across all QTL sharing fractions of about -8% on average relative to REML estimates (Fig. 2, supplementary tab. S8A, S8B). The block jackknife resampling method yielded more accurate RMSE estimates for covG CV overall (RMSE∈[0.0329,0.0365], rel.Δ∈[-12%,-14%]) but was seen to be slightly more biassed on average compared to variable block-count sampling (RMSE∈[0.0390, 0.0481], rel.Δ∈[-12%, 5%]), across block definitions and measured across all simulated traits and QTL sharing fractions. Using the number-of-variants block definition yielded slightly more accurate but more biassed covG CV estimates on average (RMSE∈[0.0329,0.0392] | rel.Δ∈[-12%, -14%]) compared to the recombination distance block definition (RMSE∈[0.0362,0.0481] | rel.Δ∈[-12%, 5%]), across (re)sampling methods and measured across all simulated traits and QTL sharing fractions.

**Weighting:** The alternate correlation correction weighting was seen to yield a tiny but consistent improvement to the accuracy when applied to the simulated data, across (re)sampling methods and measured across all simulated traits and QTL sharing fractions.

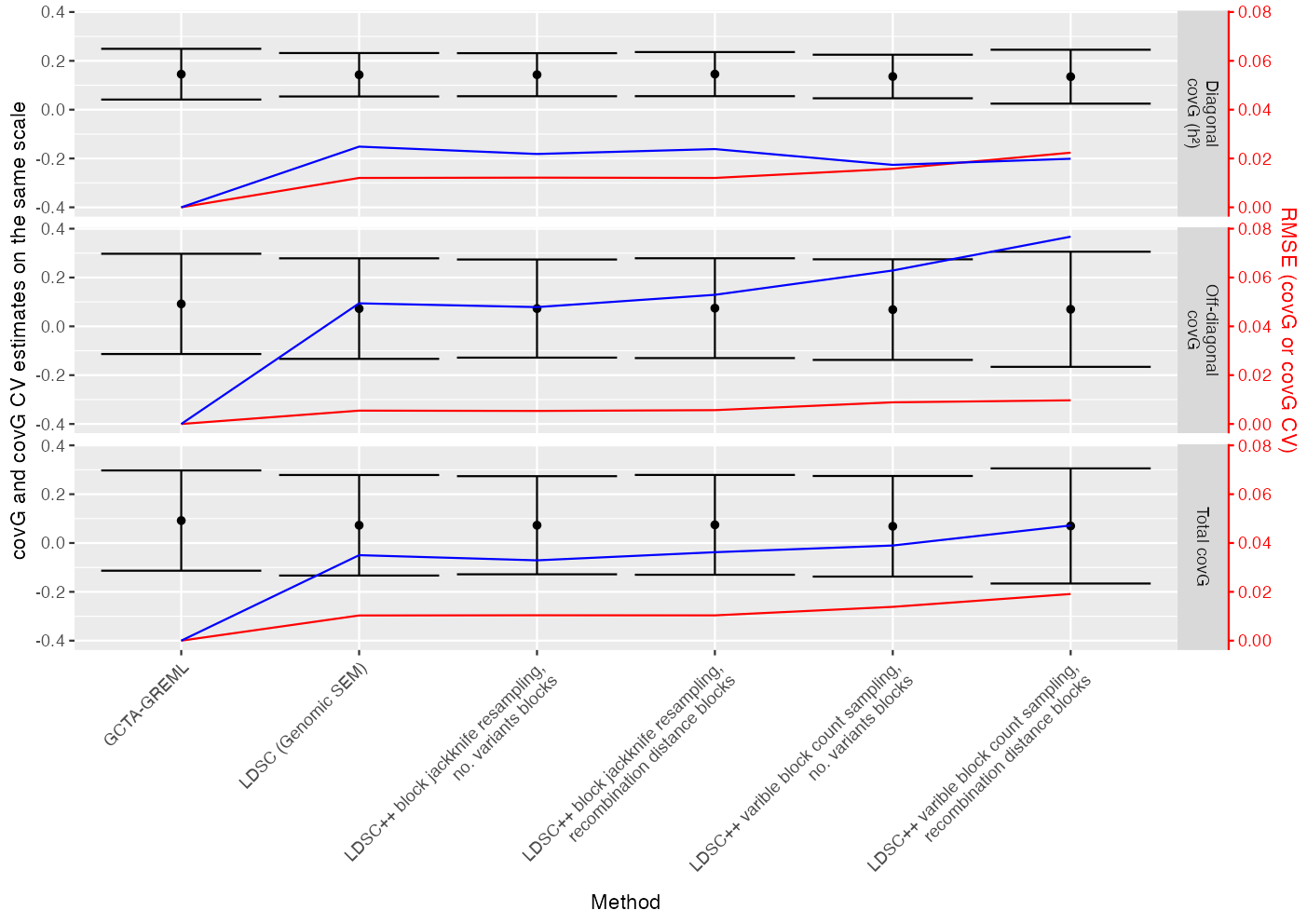

**Figure 2. Average covG with average covG CV estimates of simulated traits**

Average estimates of covG (black), separating off-diagonal covG elements, diagonal elements (h^2^), and the total average, from a genetic covariance matrix. Averages were measured over all four simulated trait pairs with either 25%, 50%, 75%, or 100% fraction of shared quantitative trait loci. Error bars (black) represent the corresponding average covG CV estimates for each covG (not confidence intervals!). RMSE deviances from GCTA-GREML estimates are shown superimposed for covG (red) and covG CV (blue), which both use the red scale on the right. Results are shown for each method along the horizontal axis. All LDSC++ method combinations use the adjusted weighting scheme to correct for correlation between the dependent and independent regression variable.

#### 2.5 Extensions to LD score regression applied to external real world GWAS data

See the main manuscript for validation results on individual level data from our GLAD+ sample.

##### 2.5.1 Extension 1, 2 & 3 - Variable block-count, extended block definitions to allow for variable block-count, and variable block-count sampling

We compared LDSR results from different combinations of (re)sampling method and block definition while allowing for a varying number of blocks (see below and supplementary tab. S9A), without activating our weighting extensions. Standard LDSR settings; block jackknife resampling and a fixed number of 200 blocks, were used as comparison reference. The default block-size setting for the number-of-variants block definition resulted in block-counts in the range of 249 - 405, without INFO filter. The default block-size setting for the recombination distance block definition resulted in block-counts in the range of 338 - 364, without INFO filter. As expected, computations for off-diagonal bivariate trait combinations obtained smaller block-counts on average due to reduced variant overlap between the trait pair GWASs and/or the reference panel (supplementary tab. S6). For the number-of-variants block definition, the lowest average block-count was seen for trait pairs including the SCHI GWAS with 249 - 269 blocks. This was expected due to the SCHI GWAS low number of variants in its summary statistics, which we estimated to 0.6 of the median dataset size after filters and merging with the reference panel list of variants. When using recombination distance blocks, this issue had a lesser influence on the block-count.

###### covG comparison results

- Number-of-variants block definition
  - Variable block-count sampling: This setting resulted in an average relative difference in covG of 1.7% or 4.4% when only considering h^2^. Individual statistically significant results reached relative differences of 11.2%, while most did not reach statistical significance. The Wilcoxon signed-rank test did indicate significant difference (p<0.001) in covG between the methods however.
  - Block jackknife resampling: Not applicable, since covG is not influenced by allowing the number of blocks to vary when using block jackknife resampling.
- Recombination distance block definition
  - Variable block-count sampling: This setting resulted in an average relative difference in covG of 0.8%, and 7.6% when only considering h^2^. Many of the differences were also statistically significant, among which were significant results for the differences in h^2^ for BIPD, BMI, MDDB, INSO, and SCHI. Individual statistically significant results reached relative differences of 23.5%. The Wilcoxon signed-rank test indicated significant difference (p<0.001) in covG between the methods.
  - Block jackknife resampling: Not applicable, since covG is not influenced by allowing the number of blocks to vary when using block jackknife resampling.

###### covG variance comparison results

- Number-of-variants block definition
  - Variable block-count sampling: This setting yielded an average difference in covG CV of -4.2%, or -3% when considering h^2^ CV only. A majority of the differences were seen to be statistically significant. Individual statistically significant results had relative differences in the range from -18.1% to 13.4%. However, the Wilcoxon signed-rank test did not indicate a significant difference in covG variance between the methods.
  - Block jackknife resampling: This setting resulted in an average relative difference in covG CV of -2.0%, or -1.3% considering h^2^ CV only. A number of these differences were seen as statistically significant in our tests. Individual statistically significant results had relative differences in the range from -8.7% to 12.3%. The Wilcoxon signed-rank test indicated significant (p=0.005) differences in covG variance between the methods.
- Recombination distance block definition
  - Variable block-count sampling: This setting yielded an average relative difference in covG CV of 8.1%, or 6.7% considering h^2^ CV only. Most of these differences were seen as statistically significant in our tests. Individual statistically significant results had relative differences in the range from -20.3% to 36.2%. The Wilcoxon signed-rank test indicated significant (p<0.001) differences in covG variance between the methods.
  - Block jackknife resampling: This setting yielded an average relative difference in covG CV of 0.5%, or 2.1% considering h^2^ CV only. A few of these differences were seen as statistically significant in our tests. Individual statistically significant results had relative differences in the range of -8.3% to 12.2%. The Wilcoxon signed-rank test did not indicate significant differences in covG variance between the methods.

##### 2.5.2 Extension 4 - Extended weighting scheme for imputation quality

We compared LDSR results from applying our weighting scheme for imputation quality (see below, supplementary tab. S9A) across different combinations of (re)sampling method and block definition while allowing for a varying number of blocks. As reference, we used LDSR with the corresponding (re)sampling method and block definition, but without weighting extensions activated to validate the weighting extensions independently. Larger differences were expected for the GWAS summary statistics that were found to include imputation quality INFO scores (Tab. 1), and included ranges with INFO scores less than ~0.8-0.9; i.e. BIPD, BMI, and MDD.

###### covG comparison results

- Number-of-variants block definition
  - Variable block-count sampling: We found a significant relative difference for BIPD h^2^ ( 5%).
  - Block jackknife resampling: We found a significant relative difference for BIPD h^2^ (4.3%).
- Recombination distance block definition
  - Variable block-count sampling: We found a significant relative difference for BIPD h^2^ (4.9%).
  - Block jackknife resampling: We found a relative difference for BIPD h^2^ (4.7%).

###### covG variance comparison results

- Number-of-variants block definition
  - Variable block-count sampling: We found a relative difference in BIPD-ANXI covG CV of -5.7% (q<0.001) and BIPD-MDDB covG CV of -5.1% (q<0.001).
  - Block jackknife resampling: We found a relative difference in BIPD-MDDB covG CV of -7.3% (q<0.001).
- Recombination distance block definition
  - Variable block-count sampling: We found significant relative differences in BIPD-ANXI covG CV of -6.5% (q<0.001), BIPD-BMI covG CV of -5.8% (q<0.013), BIPD-MDDS covG CV of -2.3% (q<0.013), and BIPD-EDUA covG CV of -0.6% (q<0.017).
  - Block jackknife resampling: We found a significant relative difference in BIPD-ANXI covG CV of -7.2% (q=0.001).

##### 2.5.3 Extension 5 - Adjusted weighting scheme to correct for the correlation between LD score and the association statistic

We compared LDSR results from applying our weighting scheme for correlation correction (see below and supplementary tab. S7) separately across different combinations of (re)sampling method and block definition while allowing for a varying number of blocks. As reference, we used LDSR with the corresponding (re)sampling method and block definition, but without weighting extensions activated to validate the weighting extensions independently.

###### covG comparison results

- Number-of-variants block definition
  - Variable block-count sampling: No individually significant results were seen. The Wilcoxon signed-rank test did show that the overall differences, with an average relative difference of -0.5% for h^2^ only, were significant however (p=0.004).
  - Block jackknife resampling: No individually significant results were seen. However, the Wilcoxon signed-rank test did show that the overall differences, with an average relative difference of -0.7% for h^2^ only, were significant (p<0.001). As expected, this result was the same across block definitions for block jackknife resampling.
- Recombination distance block definition
  - Variable block-count sampling: No individually significant results were seen. The Wilcoxon signed-rank test did show that the overall differences, with an average relative difference of -0.6% for h^2^ only, were significant however (p=0.004).
  - Block jackknife resampling: No individually significant results were seen. However, the Wilcoxon signed-rank test did show that the overall differences, with an average relative difference of -0.7% for h^2^ only, were significant (p<0.001).

###### covG variance comparison results

- Number-of-variants block definition
  - Variable block-count sampling: No individually significant results were seen. The Wilcoxon signed-rank test did show that the overall differences, with an average relative difference of 0.7% for h^2^ only, were significant however (p<0.001).
  - Block jackknife resampling: No individually significant results were seen. However, the Wilcoxon signed-rank test did show that the overall differences, with an average relative difference of 1.2% for h^2^ only, were significant (p<0.001).
- Recombination distance block definition
  - Variable block-count sampling: No individually significant results were seen. The Wilcoxon signed-rank test did show that the overall differences, with an average relative difference of 0.7% for h^2^ only, were significant however (p<0.001).
  - Block jackknife resampling: No individually significant results were seen. The Wilcoxon signed-rank test did show that the overall differences, with an average relative difference of 1.3% for h^2^ only, were significant however (p<0.001).

##### 2.5.4 Combined extensions

We compared LDSR results from using either combination of (re)sampling method and block definition while simultaneously applying both extended weighting schemes and allowing for a varying number of blocks (supplementary tab. S9A). Standard LDSR settings; block jackknife resampling and a fixed number of 200 blocks, were used as comparison reference.

###### covG comparison results

- Number-of-variants block definition
  - Variable block-count sampling: This setting resulted in an average relative difference in covG of 1.4% or 4.3% when only considering h^2^. A number of the individual covG differences were statistically significant, which were ANXD-BIPD (10.3%), BIPD (12.1%), INSO (4.5%), and SCHI (4.7%). The Wilcoxon signed-rank test indicated a significant difference (p<0.001) in covG between the methods.
  - Block jackknife resampling: This setting resulted in an average relative difference in covG of -0.2% or -0.3% when only considering h^2^. No individual significant covG differences were seen. The Wilcoxon signed-rank test indicated a significant difference (p=0.01) in covG between the methods, however.
- Recombination distance block definition
  - Variable block-count sampling: This setting resulted in an average relative difference in covG of -1%, and 7.3% when only considering h^2^. Most of the differences were also statistically significant. Individual statistically significant results had relative differences in the range of -72.2 - 23.1%. The Wilcoxon signed-rank test indicated a significant difference (p<0.001) in covG between the methods.
  - Block jackknife resampling: This setting resulted in an average relative difference in covG of -0.6% (equally when only considering h^2^). No individual significant covG differences were seen. The Wilcoxon signed-rank test indicated a significant difference (p=0.01) in covG between the methods, however.

###### covG variance comparison results

- Number-of-variants block definition
  - Variable block-count sampling: This setting yielded an average difference in covG CV of -4.2%, or -2.7% when considering h^2^ CV only. Most of the differences were seen to be statistically significant. Individual statistically significant results had relative differences in the range of -17.5 - 13.9%. However, the Wilcoxon signed-rank test did not indicate a significant difference in covG variance between the methods.
  - Block jackknife resampling: This setting resulted in an average relative difference in covG CV of -1.6%, or -.3% considering h^2^ CV only. Most of these differences were seen as statistically significant in our tests. Individual statistically significant results had relative differences in the range of -10.1 - 13.2%. The Wilcoxon signed-rank test indicated a significant (p=0.034) difference in covG variance between the methods.
- Recombination distance block definition
  - Variable block-count sampling: This setting yielded an average relative difference in covG CV of 8.3%, or 7.2% considering h^2^ CV only. Most of the individual differences were seen as statistically significant in our tests. Individual statistically significant results had relative differences in the range of -24.8 - 36.6%. The Wilcoxon signed-rank test indicated a significant (p<0.001) difference in covG variance between the methods.
  - Block jackknife resampling: This setting yielded an average relative difference in covG CV of -1.0%, or 3.1% considering h^2^ CV only. Many of the individual differences were seen as statistically significant in our tests. Individual statistically significant results had relative differences in the range of -1.9 - 13.3%. The Wilcoxon signed-rank test did not indicate a significant difference in covG variance between the methods, however.
